# Comprehensive mapping of neutralizing antibodies against SARS-CoV-2 variants induced by natural infection or vaccination

**DOI:** 10.1101/2021.05.03.21256506

**Authors:** Xinhua Chen, Zhiyuan Chen, Andrew S. Azman, Ruijia Sun, Wanying Lu, Nan Zheng, Jiaxin Zhou, Qianhui Wu, Xiaowei Deng, Zeyao Zhao, Xinghui Chen, Shijia Ge, Juan Yang, Daniel T. Leung, Hongjie Yu

## Abstract

**Background:** Immunity after SARS-CoV-2 infection or vaccination has been threatened by recently emerged SARS-CoV-2 variants. A systematic summary of the landscape of neutralizing antibodies against emerging variants is needed.

**Methods:** We systematically searched PubMed, Embase, Web of Science, and 3 pre-print servers for studies that evaluated neutralizing antibodies titers induced by previous infection or vaccination against SARS-CoV-2 variants and comprehensively collected individual data. We calculated lineage-specific GMTs across different study participants and types of neutralization assays.

**Findings:** We identified 56 studies, including 2,483 individuals and 8,590 neutralization tests, meeting the eligibility criteria. Compared with lineage B, we estimate a 1.5-fold (95% CI: 1.0-2.2) reduction in neutralization against the B.1.1.7, 8.7-fold (95% CI: 6.5-11.7) reduction against B.1.351 and 5.0-fold (95% CI: 4.0-6.2) reduction against P.1. The estimated neutralization reductions for B.1.351 compared to lineage B were 240.2-fold (95% CI: 124.0-465.6) reduction for non-replicating vector platform, 4.6-fold (95% CI: 4.0-5.2) reduction for RNA platform, and 1.6-fold (95% CI: 1.2-2.1) reduction for protein subunit platform. The neutralizing antibodies induced by administration of inactivated vaccines and mRNA vaccines against lineage P.1 were also remarkably reduced by an average of 5.9-fold (95% CI: 3.7-9.3) and 1.5-fold (95% CI: 1.2-1.9).

**Interpretation:** Our findings indicate that the antibody response established by natural infection or vaccination might be able to effectively neutralize B.1.1.7, but neutralizing titers against B.1.351 and P.1 suffered large reductions. Standardized protocols for neutralization assays, as well as updating immune-based prevention and treatment, are needed.

**Funding:** Chinese National Science Fund for Distinguished Young Scholars

**Research in context:** *Evidence before this study:* Several newly emerged SARS-CoV-2 variants have raised significant concerns globally, and there is concern that SARS-CoV-2 variants can evade immune responses that are based on the prototype strain. It is not known to what extent do emerging SARS-CoV-2 variants escape the immune response induced by previous infection or vaccination. However, existing studies of neutralizing potency against SARS-CoV-2 variants are based on limited numbers of samples and lack comparability between different laboratory methods. Furthermore, there are no studies providing whole picture of neutralizing antibodies induced by prior infections or vaccination against emerging variants. Therefore, we systematically reviewed and quantitively synthesized evidence on the degree to which antibodies from previous SARS-CoV-2 infection or vaccination effectively neutralize variants.

*Added value of this study:* In this study, 56 studies, including 2,483 individuals and 8,590 neutralization tests, were identified. Antibodies from natural infection or vaccination are likely to effectively neutralize B.1.1.7, but neutralizing titers against B.1.351 and P.1 suffered large reductions. Lineage B.1.351 escaped natural-infection-mediated neutralization the most, with GMT of 79.2 (95% CI: 68.5-91.6), while neutralizing antibody titers against the B.1.1.7 variant were largely preserved (254.6, 95% CI: 214.1-302.8). Compared with lineage B, we estimate a 1.5-fold (95% CI: 1.0-2.2) reduction in neutralization against the B.1.1.7, 8.7-fold (95% CI: 6.5-11.7) reduction against B.1.351 and 5.0-fold (95% CI: 4.0-6.2) reduction against P.1. The neutralizing antibody response after vaccinating with non-replicating vector vaccines against lineage B.1.351 was worse than responses elicited by vaccines on other platforms, with levels lower than that of individuals who were previously infected. The neutralizing antibodies induced by administration of inactivated vaccines and mRNA vaccines against lineage P.1 were also remarkably reduced by an average of 5.9-fold (95% CI: 3.7-9.3) and 1.5-fold (95% CI: 1.2-1.9).

*Implications of all the available evidence:* Our findings indicate that antibodies from natural infection of the parent lineage of SARS-CoV-2 or vaccination may be less able to neutralize some emerging variants, and antibody-based therapies may need to be updated. Furthermore, standardized protocols for neutralizing antibody testing against SARS-CoV-2 are needed to reduce lab-to-lab variations, thus facilitating comparability and interpretability across studies.

## Introduction

Since the first sequence of SARS-CoV-2 was published in January of 2020^1^, over 1.1 million strains have been documented in Global Influenza Surveillance and Response System (GISAID)^2^, with recent reports of several newly emerged lineages, which has raised significant concerns globally. Of particular concern has been the emergence of lineage B.1.1.7 (UK variant, also known as 501Y.V1), lineage B.1.351 (South Africa variants, 501Y.V2), and lineage P.1 (Brazil variant, 501Y.V3), harboring several significant mutations in spike glycoproteins, which are key domains of virus-neutralizing antibodies^3^. These mutants rapidly became the dominant circulating virus strains in the regions where they were first isolated, and especially for B.1.1.7 and B.1.351, spread globally. Several vital mutations in the receptor-binding domain (RBD), such as N501Y in B.1.1.7 and E484K in B.1.351 and P.1, are associated with increased infectivity and decreased neutralizing potency, with potential to evade humoral immunity from prior infections or vaccinations^4-8^. Another variant of lineage B.1.427/B.1.429, which first emerged in California, was categorized as a Variant of Concern (VOC) in March 2021 according to classification developed by SARS-CoV-2 Interagency Group (SIG) in United States, containing a single L452R mutation in RBD in spike, whose ability of reduced sensitivity to neutralization has yet to be determined^9^.

There is concern that SARS-CoV-2 variants can evade immune responses elicited by natural infections and vaccines that are based on the prototype strain (Wuhan-Hu-1). It has been shown that neutralization against variants by convalescent plasma is remarkably reduced by several mutations, including E484K shared by lineage B.1.351 and P.1^10^. Serum collected from recipients of licensed vaccines have a decreased ability to neutralize emerging SARS-CoV-2 mutants to varying degrees, from 1.6-fold reduction for China’s protein subunit vaccines to over 6-fold reduction for mRNA vaccines^11-15^. Additionally, consistent with immunogenicity results, a major loss of efficacy against B.1.351 was seen in NVX-CoV2373 and ChAdOx1 nCoV-19 vaccines^16,17^, though the efficacy was retained against B.1.1.7 for other vaccines^18^. However, existing studies of neutralizing potency against SARS-CoV-2 variants are based on limited numbers of samples and lack comparability between different laboratory methods. Furthermore, there are no studies providing whole picture of neutralizing antibodies induced by prior infections or vaccination against emerging variants.

Here, we systematically summarize the evidence on neutralization ability against various SARS-CoV-2 variants among those previously-infected with strains from the original SARS-CoV-2 lineage and those who have been vaccinated.

## Methods

### Study Selection and Data Extraction

We conducted a systematic search from six databases, including three peer-reviewed databases (PubMed, Embase and Web of Science) and three preprint servers (medRxiv, bioRxiv and Europe PMC), for studies published in English between Sep 1, 2020 and Apr 18, 2021 with predefined search terms (**Table S1**). We included studies that 1) reported neutralizing antibodies against SARS-CoV-2 variants by using serum or plasma collected from individuals with virologically or serologically-confirmed SARS-CoV-2 infections, and vaccine recipients; and 2) reported or displayed individual antibody titers with summary tables or high-resolution images. Studies that 1) investigated the efficacy of monoclonal and therapeutic antibodies against variants; 2) reported seroprevalence of variants; 3) only detected non-neutralizing antibodies; or 4) reported specific mutation sites from a view of biological mechanism, were excluded. We also excluded abstracts of congress meetings or conference proceedings, study protocols, media news, commentaries, and reviews.

We screened all eligible studies to extract the study characteristics, study participants, types of variants, laboratory methods and antibody titers (**Table S2**). Data were digitized from the figures in papers by pre-trained investigators with a digital extraction tool if individual titer values were not available in table format^19^. We only extracted the titers expressed as reciprocal dilution of serum that neutralizes or inhibits 50% of the virus (e.g., NT50, PRNT50, etc.). When titers were not explicitly stated, but a category defined as less than some value exists, we assumed a titer of half of this value (e.g., titer of 10 is assumed when “<20” was present). For individuals with multiple specimens, we only included one sample that most likely to have neutralization antibodies for each study participant to avoid repeated inclusion. The inclusion and exclusion of studies, screening and scrutinization of included studies, data extraction and verification were performed by two independent researchers, a third researcher was consulted when disagreement arose.

### Data Synthesis and Analysis

The primary outcome variable was a pooled geometric mean titer (GMT), expressing an average level SARS-CoV-2 neutralizing titers for a group of individual titers. To be specific, we calculated lineage-specific GMT with extracted dataset across different study participants and types of neutralization assays, indicating three stratified factors (i.e., study participants, types of neutralization assays and lineages).

Individual-level data were classified into four groups (**Table S3**). Briefly, 1) non-variant-infected individuals, which indicates acutely-infected or convalescent COVID-19 patients infected with parental strains or asymptomatic cases infected with non-variants; 2) variant-infected individuals, which refers to individuals infected with SARS-CoV-2 variants; 3) uninfected vaccine recipients, which refers to healthy vaccines who were not infected with either parental strains or the variants of SARS-CoV-2; 4) previously-infected vaccine recipients, which refers to vaccines who had been infected with parental strains.

Among different study participants, we further stratified studies by types of neutralization assays. Multiple neutralization assays used in included studies were classified into three categories based on the type of virus (authentic or pseudo) used and the types of vectors [lentivirus or vesicular stomatitis virus (VSV)] used in pseudovirus neutralization assays, namely, live virus neutralization assays, lentivirus-vector pseudovirus neutralization assays, and VSV-vector pseudovirus neutralization assays.

Within specific study participants and types of neutralization assays, we calculated lineage-specific GMTs. The categories of SARS-CoV-2-variant lineages among included studies was consistent with taxonomy and classification of Phylogenetic Assignment of Named Global Outbreak Lineage (PANGO lineage)^20^. Specifically, lineage B.1.1.117, B.1.1.26, B.1.1.50, and B.1.1.29 served as the reference strains when comparing with SARS-CoV-2 variants in some studies^11,16,21,22^, and they were classified as lineage B.1 due to close phylogenetic distance and shared mutation site of 614G^22,23^. Emerging and circulating SARS-CoV-2 variants have been divided into three classes by SIG: variant of interest (VOI), variant of concern (VOC), and variant of high consequence (VOHC)^24^. VOIs include lineage B.1.526, B.1.525 and P.2, with increased transmissibility and disease severity, and VOCs include lineage B.1.1.7, B.1.351, P.1, B.1.427, and B.1.429, with significant reduction in neutralization and increased hospitalizations or deaths^24^. At the time of writing, no variants have been classified as VOHC^24^. The classification of all lineages involved in eligible studies were shown in **Table S4**.

For non-variant-infected subjects, we further explored potential determinants affecting the GMT, such as the sampling interval post symptom onset, and disease severity. Disease severity was assessed by classifying individuals as either hospitalized or non-hospitalized, based on World Health Organization COVID-19 Clinical management criteria^25^. We divided the post-symptom-onset sampling interval into three periods (i.e., 0-30 days, 31-90 days, and >90 days) according to the distribution of sampling times in the extracted data (**Figure S9**). We stratified the uninfected vaccine recipients by vaccine platforms (e.g., non-replicating vector, RNA, inactivated, and subunit protein) and the sampling interval post vaccination (i.e., <14 days, 14-90 days, and >90 days) based on a study that evaluated antibody persistence through 6 months after vaccination^26^. Furthermore, we conducted a matched analysis for samples that had been tested simultaneously against reference strains and variants and calculated the fold changes of GMT.

### Statistical Analysis

We first performed univariate subgroup analysis, then used multivariate linear regression models to control for potential confounding among non-variant-infected individuals and uninfected vaccinees to quantitatively explore potential factors that may affect GMT. For the multivariate regression model, we included pango lineage, vaccine platform, sampling interval after last dose, and sampling intervals post symptom onset, and disease severity with available data. GMT was calculated as arithmetic mean of log-transformed titer based on natural logarithm form, and t-test 95% CI was estimated. Statistical significance was tested by Kruskal-Wallis rank sum test with Nemenyi’s post-hoc test. For paired samples, we used Wilcoxon matched-pairs signed-rank test. Only P-value less than 0.05 were considered statistically significant (P ≤ 0.05, *; P ≤ 0.01, **; P ≤ 0.001, ***). All statistical analyses were done using R (version 4.0.1).

## Results

### Study selection and data extraction

We identified a total of 5,182 studies after systematically searching multiple data sources with 2,307 coming from peer-reviewed databases, and 2,875 from preprint severs (**Figure 1**). After screening title, abstract, and full-text, 56 studies containing a total of 2,483 individuals and 8,590 neutralization measurements were included in our analysis, with previously uninfected vaccine recipients comprising more than half of studies (44 studies; 4,697/8,590 samples, 54.7%), followed by non-variant-infected individuals (41 studies; 3,440/8,590 samples, 40.0%), variant-infected individuals (8 studies; 288/8,590 samples, 3.4%), and previously-infected vaccine recipients (6 studies; 165/8,590 samples, 1.9%) (**Figure 1**). Live virus neutralization assays were most common (24 studies, 24/56, 42.9%), followed by lentivirus-vector pseudovirus neutralization assay (22 studies, 22/56, 39.3%) and VSV-vector pseudovirus neutralization assay (13 studies, 13/56, 23.2%) (**Table S7)**. The lineage B.1.1.7 and B.1.351 were the two SARS-CoV-2 variants that had been studied the most, comprising more than two third of studies (B.1.1.7: 37 studies, 66.1%; B.1.351: 40 studies, 71.4%) and nearly half of measurements (B.1.1.7: 1,852 measurements, 21.6%; B.1.351: 2,422 measurements, 28.2%) (**Table S7**). Other VOCs, such as lineage P.1 and B.1.427/B.1.429 were the subject of 20 studies including 863 data points (**Table S7**).

**Figure 1.**
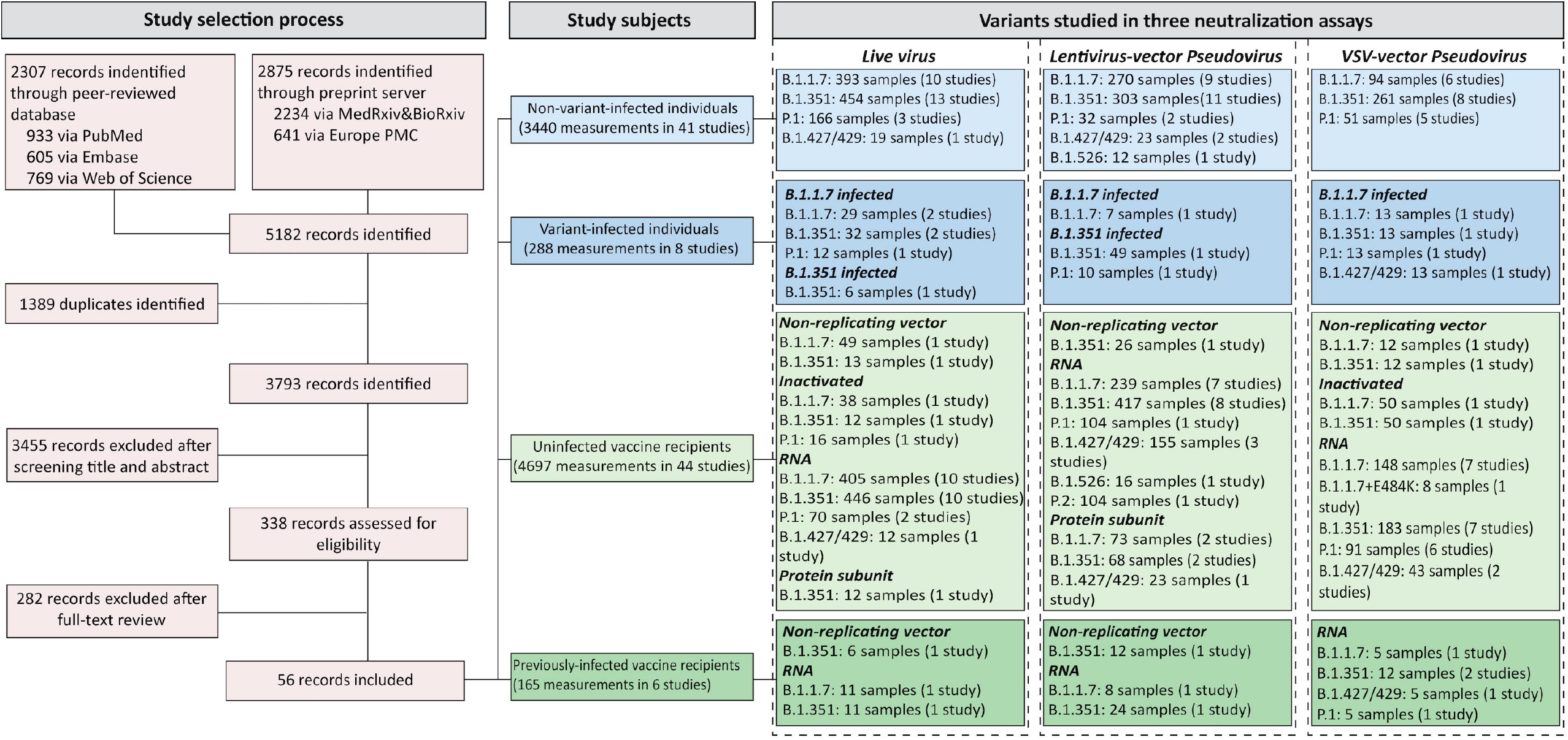
Selection flowchart of studies, study participants, and variants studied.

### Level of neutralizing antibodies against SARS-CoV-2 variants among non-variant-infected individuals

Overall, among studies that evaluated neutralizing antibodies in individuals previously infected with nonvariants, 26.3% (5/19), 95.7% (22/23), and 88.9% (8/9) of the studies found a significant decrease in neutralization against B.1.1.7, B.1.351 and P.1, respectively. The neutralization levels of 6.2% (47/757), 20.0% (204/1,028), and 6.4% (16/249) samples against B.1.1.7, B.1.351 and P.1 were reduced to below the limitation of detection.

In aggregate, neutralizing titers against B.1.351 were significantly reduced, followed by P.1, while titers against B.1.17 were not, when compared to reference lineages titers. With live virus neutralization assays, the pooled GMT was 254.6 (95% CI: 214.1-302.8) for lineage B.1.1.7, 79.2 (95% CI: 68.5-91.6) for B.1.351, 253.1 (95% CI: 194.9-328.8) for P.1, and 275.5 (95% CI: 146.4-518.5) for B.1.427/B.1.429, with an average of 3.4-fold (95% CI: 3.0-4.0) in B.1.351 when comparing to lineage B (**Figure 2A**). The decrease in neutralization for B.1.351 (3.5, 95% CI: 3.0-4.0) is similar when compared to lineage B.1, which has the 614G mutation. In matched analyses, the reduction in the ability of neutralization against B.1.1.7, B.1.351 and P.1 was 1.5 (95% CI: 1.0-2.2), 8.7 (95% CI: 6.5-11.7) and 5.0-fold (95% CI: 4.0-6.2) compared with lineage B (**Figure S2**). For lentivirus-vector pseudovirus assays, the average reductions of GMT were 8.1-9.7 fold, 4.2-5.1 fold, and 1.4-1.6 fold for lineage B.1.351, P.1, and B.1.1.7 compared to lineage B/B.1/B+D614G (**Figure 2B**). In terms of VSV-vector pseudovirus assays, the average reductions of GMT were 2.0-12.3 fold, 1.2-7.9 fold, and 1.2-7.5 fold for lineage B.1.351, P.1, and B.1.1.7 respectively when compared to reference strains (**Figure 2C**).

**Figure 2.**
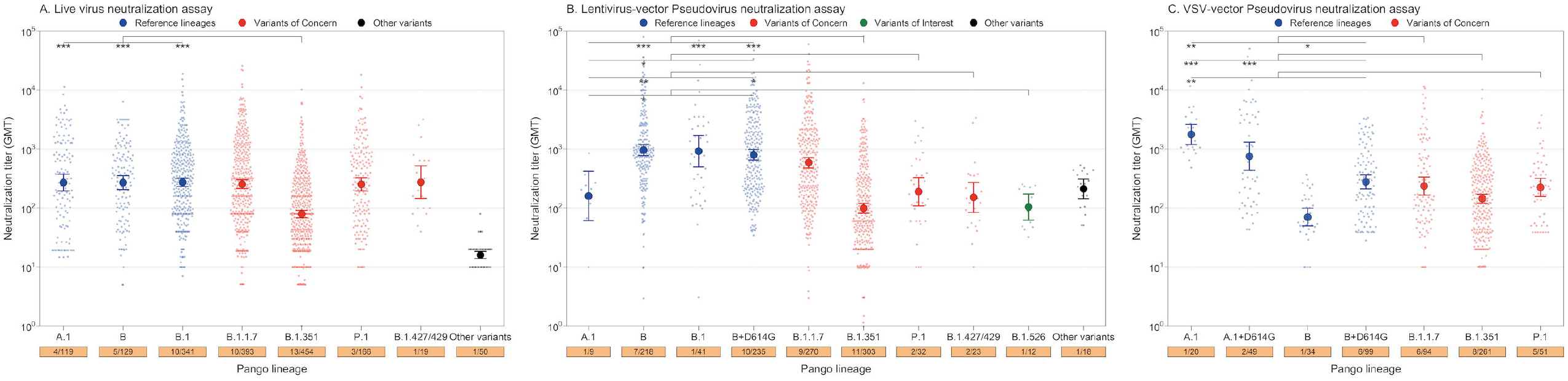
Neutralizing antibodies against SARS-CoV-2 variants in non-variant-infected individuals. Neutralizing antibodies against reference strains (blue dot) and variants of concern (red dot) were determined in **A)** live virus neutralization assay, **B)** lentivirus-vector pseudovirus neutralization assay, and **C)** VSV-vector pseudovirus neutralization assay. The solid point represents the GMT and the error bar represents the 95% confidence interval. The scattering dot represents individual titers. The numbers in the bottom orange rectangle represent the number of studies and sample sizes (no. of studies/no. of samples). Significant statistical differences are indicated by asterisks (P ≤ 0.05, *; P ≤ 0.01, **; P ≤ 0.001, ***).

Serum collected from individuals infected with SARS-CoV-2 variants show increased neutralizing antibodies level against corresponding mutant strains (**Figure S3**). Notably, B.1.1.7-infected persons also show significant decreased neutralizing antibodies titers against B.1.351 (**Figure S3**). Multivariate analysis also indicated B.1.351had significantly reduced neutralizing activity for all three neutralization assays after controlling for sampling intervals post symptom onset and/or clinical severity, compared to reference lineages (**Table S8**).

### Level of neutralizing antibodies against SARS-CoV-2 variants among vaccine recipients

Significant reductions in neutralization against B.1.1.7, B.1.351 and P.1 were found in 48.1% (13/27), 100.0% (26/26) and 66.7% (6/9) of the studies in SARS-CoV-2 naïve vaccine recipients, respectively. Partial reductions reached the background levels, with the proportion of 4.0 (41/1,014), 19.1 (237/1,239), and 11.3 (32/281) against B.1.1.7, B.1.351 and P.1, respectively.

Serum collected from uninfected vaccine recipients had diverse neutralizing antibody levels against SARS-CoV-2 variants across different vaccine platforms. In studies using the live virus neutralization assay, the non-replicating adenoviral vectored vaccine showed generally low GMT against B.1.351 in contrast to other vaccine delivery platforms, with an average of GMTs of 2.1 (95% CI: 1.1-4.1), 70.9 (95% CI: 50.8-98.9), 85.9 (95% CI: 75.9-97.2), and 66.6 (95% CI: 51.0-86.9) for platforms of non-replicating vector, inactivated virus, RNA, and protein subunit, respectively (**Figure 3A**). Using lineage B as a reference lineage, we found that the average reduction fold of neutralizing antibodies against B.1.351 was 240.2-fold (95% CI: 124.0-465.6) for non-replicating vector platform, 4.6-fold (95% CI: 4.0-5.2) for RNA platform, and 1.6-fold (95% CI: 1.2-2.1) for protein subunit platform (**Figure 3A**). The neutralizing antibodies induced by administration of inactivated vaccines and mRNA vaccines against lineage P.1 were also remarkably reduced by an average of 5.9-fold (95% CI: 3.7-9.3) and 1.5-fold (95% CI: 1.2-1.9) (**Figure 3A**).

**Figure 3.**
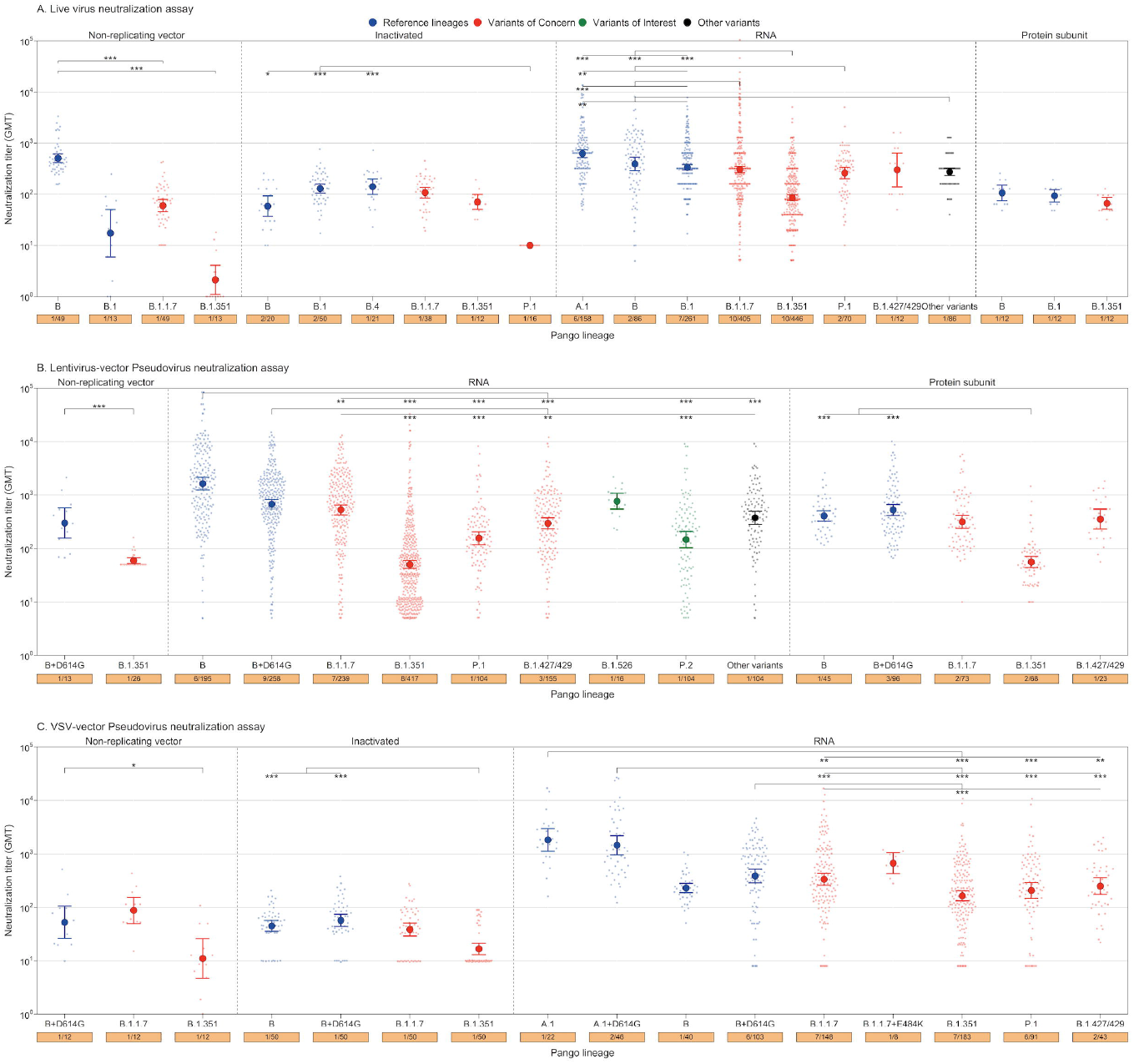
Neutralizing antibodies against SARS-CoV-2 variants in uninfected vaccine recipients after the administration of different vaccine platforms. Neutralizing antibodies against reference strains (blue dot), variants of concern (red dot), variants of interest (green dot), and other variants (black dot) were determined in **A)** live virus neutralization assay, **B)** lentivirus-vector pseudovirus neutralization assay, and **C)** VSV-vector pseudovirus neutralization assay. The solid point represents the geometric mean titer (GMT) and the error bar represents the 95% confidence interval. The scattering dot represents individual titers. The numbers in the bottom orange rectangle represent the number of studies and sample sizes (no. of studies/no. of samples). Significant statistical differences are indicated by asterisks (P ≤ 0.05, *; P ≤ 0.01, **; P ≤ 0.001, ***).

In paired-sample analysis, neutralizing titers induced by mRNA vaccine were reduced 3.1-fold (95% CI: 2.4-3.9) against B.1.1.7, 2.7-fold (95% CI: 2.3-3.2) against P.1, and 7.4-fold (95% CI: 6.4-8.5) against B.1.351, compared to prototype strain Wuhan-Hu-1 that was used for vaccine design (**Figure S4**). In comparison to lineage B+D614G, significant reductions of antibody levels elicited by all included vaccine platforms against lineage B.1.351 were also found in two pseudovirus neutralization assays (P<0.05). Neutralizing antibodies against lentiviral-encapsulated P.1, P.2, and B.1.427/B.1.429 were also significantly reduced when comparing to lineage B+614G (P<0.01) (**Figure 3B**). Additionally, antibodies in serum collected from previously-infected vaccine recipients receiving RNA vaccines or non-replicating-vector vaccines were significantly higher than uninfected vaccine recipients in pseudovirus assays (**Figure S7**). In multivariate regression mode, we found that B.1.351, P.1 and B.1.427/1.429 had significant lower GMT of neutralizing antibodies assessed by live virus neutralization assay, and we found that other vaccine platform shows significant higher antibody level than non-replicating-vector vaccine across all included virus strains (**Table S9**).

### Comparison between GMT of neutralizing antibodies induced by natural infection and vaccination

Comparisons of the GMT of neutralizing antibodies induced by natural infections (non-variant-infected individuals) and vaccination (uninfected vaccine recipients) showed that individuals infected with parental strains had similar antibodies titers with individuals vaccinated with mRNA vaccines, but significantly higher antibodies titers than individuals administrated with non-replicating vector vaccines against lineage B.1, B.1.1.7 and B.1.351, when using live virus neutralization assay to detect neutralizing antibodies (**Figure 4A**). No significant differences between titers were found between non-variant-infected individuals and RNA vaccine-recipients against B.1.1.7, B.1.351 and P.1 tested by VSV-based pseudovirus neutralization assays (**Figure 4C**).

**Fig 4.**
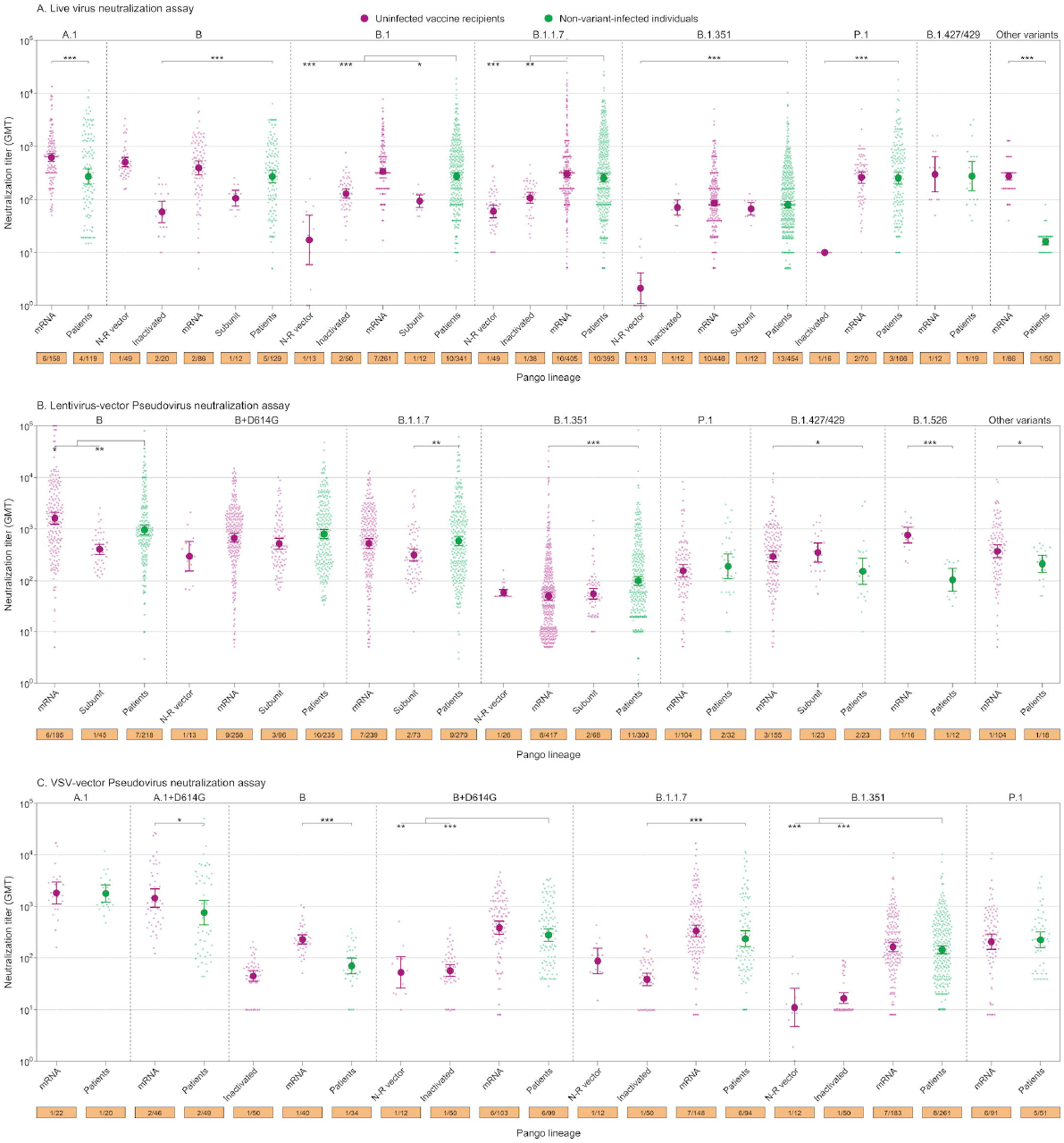
Comparison between GMT of neutralizing antibodies induced by natural infection and vaccination by platforms. Neutralizing antibodies comparison were determined in **A)** live virus neutralization assay, **B)** lentivirus-vector pseudovirus neutralization assay, and **C)** VSV-vector pseudovirus neutralization assay. Neutralization against same lineage induced by natural infection and different platforms of vaccination was compared. The solid point represents the GMT and the error bar represents the 95% confidence interval. The scattering dot represents individual titers. The numbers in the bottom orange rectangle represent the number of studies and sample sizes (no. of studies/no. of samples). Significant statistical differences are indicated by asterisks (P ≤ 0.05, *; P ≤ 0.01, **; P ≤ 0.001, ***). N-R vector, non-replicating vector.

## Discussion

Overall, we comprehensively estimate the antibody levels against preexisting SARS-CoV-2 viruses and recently emerging variants among different study participants using neutralizing antibodies as a protective biomarker. Our analyses found that antibodies in both naturally-infected and vaccine-immune sera/plasma had slightly reduced but largely retained neutralizing activity against B.1.1.7. The neutralizing potency against B.1.351 and P.1 was significantly reduced compared to reference lineages. The antibody response after vaccinating with non-replicating vector vaccines against lineage B.1.351 was worse than responses elicited by vaccines on other platforms, and the level of neutralizing antibodies is lower than that of individuals who were previously infected. This finding suggests that immunity derived from natural infection or vaccination might be less able to neutralize some recently emerging variants, and antibody-based therapies may need to be updated.

Neutralizing antibodies titers induced from both natural infections and vaccination against B.1.351 and P.1 were significantly lower than that against other variants or reference strains, mainly due to mutations in spike protein that were associated with decreased neutralizing potency to evade humoral immunity (e.g., E484K) ^27^. However, the decrease of neutralizing potency was more obvious in B.1351 than P.1, although both variants shared the E484K mutation, which could be partly explained by the distinct set of other mutations and/or deletions in the NTD region or enhanced neutralization of P.1 by anti-RBD antibodies that bind outside of the RBD^28^. We found that convalescent and vaccine-induced immune sera had neutralizing potency against B.1.1.7 and B.1.427/ B.1.429 that are comparable with reference strains, suggesting that those mutations do not remarkably affect neutralizing activity. While mutations at 501 can increase affinity for angiotensin converting enzyme 2 (ACE2) and enhance transmissibility of B.1.1.7^29-31^, variants of B.1.427/B.1.429 bearing spike mutations L452R, S13I, and W152C were only associated with modest increases in secondary household attack rates, while no evidence of reduced neutralization capacity for these L452R SARS-CoV-2 variants were found^32^.However, continued vigilance is warranted given the potential for further mutations that might affect the immunogenicity of the vaccines or reduce the cross-reactivity of previously-induced antibodies by natural infections.

Both previous infection and vaccination have been shown to provide potent protection against similar strains, but it is unclear how neutralizing antibodies against variants induced by natural infection and vaccination might be different. For the other two most prevalent variants, B.1.1.7 and B.1.351, natural infection-induced sera/plasma had significantly higher neutralizing levels than that attained by those vaccinated with non-replicating vector vaccine in live virus neutralization assay. Additionally, antibodies induced by RNA-vaccine had similar neutralizing level with antibodies derived from naturally-infected individuals against prevalent variants. This suggests that the neutralizing potency elicited by natural infection of previous prototype strains is relatively robust, whereas the immunity induced by vaccination depends on vaccine platform.

Although some studies have shown that the neutralization levels from live virus and pseudovirus correlate well,^33-35^ we stratified the results by each of three neutralization assays to enhance comparability. The results from live virus assays could provide a comprehensive way to assess inherent viral fitness and the potential impact of other mutations outside the spike region^6,33-35^. However, even within the same live virus neutralization test, different methods were used for experimental endpoints (e.g., cytopathic effect, fluorescence, etc.), and to report their final individual titers (e.g., NT50, NT80, etc.,). In addition, experimental procedures, such as virus titration, serum dilution, virus-serum neutralization, varied greatly across reports from different laboratories. Thus far, there is no integrated or standardized operation procedure for SARS-CoV-2 neutralization assay (neither live virus nor pseudovirus), making comparison between studies difficult. International efforts to standardize laboratory methods for SARS-CoV-2 neutralization tests are urgently needed. The previous work by WHO in standardizing procedures for avian influenza neutralization assays provides one useful example.^36,37^

Our study has several limitations. First, this study synthesized different neutralization assay results to estimate pooled GMTs. While we stratified analyses by neutralization assays according to virus types and vectors to get the most comparable results, significant variation between assays persists. Second, in many studies, individual-level details were not reported, thus limiting our ability to adjust for potential confounding factors in multivariate regression. We tried to contact study authors, but the response rate was generally low. Thirdly, we did not assess the impact of preexisting cellular immunity, which could provide a degree of protection as reported in previous studies^38^.

In conclusion, our study provides a comprehensive mapping of the neutralizing potency against SARS-CoV-2 variants induced by natural infection or vaccination. Our findings suggest that immune sera/plasma retained most of its neutralizing potency against B.1.1.7 and B.1.427/ B.1.429 variants, but significantly lost neutralizing potency against B.1.351 and P.1 variants, with B.1.351 having the worst reductions. The evolution of SARS-CoV-2 lineage is still in process, and it’s unknown whether long-term accumulation of mutations can erode the neutralizing effectiveness of natural and vaccine elicited immunity, especially in the context of waning immunity. Therefore, longitudinal monitoring of emerging variants and antibody-induced immunity is of high importance, and standardized protocols for neutralizing antibody testing against SARS-CoV-2 are urgently needed.

## Data Availability

Data are available upon request.

## Contributors

H.Y. designed and supervised the study. X.C. and Z.C. did the literature search, set up the database and did all statistical analyses. X.C., Z.C., A.S.A., and D.T.L. co-drafted the first version of the article. X.C., Z.C., R.S., W.L., N.Z., J.Z., Q.W., X.D., Z.Z., X.C, S.G., and helped with checking data and did the figures. D.T.L., A.S.A., J.Y, and H.Y. commented on the data and its interpretation, revised the content critically. All authors contributed to review and revision and approved the final manuscript as submitted and agree to be accountable for all aspects of the work.

## Acknowledgments

We thank Junbo Chen, Qianli Wang, and Yuxia Liang from the Fudan University. This study was funded by the National Science Fund for Distinguished Young Scholars (grant no. 81525023), Program of Shanghai Academic/Technology Research Leader (grant no. 18XD1400300), National Science and Technology Major project of China (grant no. 2018ZX10713001-007, 2017ZX10103009-005, 2018ZX10201001-010), the US National Institutes of Health (R01 AI135115 to D.T.L. and A.S.A.)

