## Appendix for "Comprehensive mapping of neutralizing antibodies against SARS-CoV-2 variants induced by natural infection or vaccination"

### Methods and results

*Data extraction and variable list*

We screened all eligible studies to extract the following information: 1) literature information, 2) study participants, 3) types of variants; 4) laboratory methods; 5) predefined outcomes, i.e., antibody titer or geometric mean titer.

The listed variables were extracted from qualified studies, including the first author’s name, title, published journal or preprints severs, publication date, study participants (vaccine recipients or previously-infected individuals), sampling time, study location, age, vaccine platform, vaccine name and doses for vaccine recipients, clinical severity and underlying conditions for COVID-19 patients. Variants’ lineages, virus names and mutation profiles were also extracted for each included study. If such information were not reported by the author, additional sources, such as Global Influenza Surveillance and Response System (GISAID, <https://www.gisaid.org/>) and Phylogenetic Assignment of Named Global Outbreak Lineage (PANGO lineages) (<https://cov-lineages.org/>) served as supplements. The laboratory methods mainly included the neutralization assays used, tested antibodies, sample type (sera or plasma), and other factors that may affect the serological results, such as dilution factor, number of duplicates, cell line, virus titer used in the assay. We recorded the individual antibody titers if the data reported by the studies, otherwise digitized the data with sufficient detail from the figures in the studies if it were not reported.

*Categories of lineages of SARS-CoV-2 variants*

The categories of SARS-CoV-2-variants lineages in this study was consistent with taxonomy and classification of PANGO lineage. Some studies had focused on significant mutation sites or their combinations (e.g., D614G+501Y+69-70del), and these mutant viruses would be classified as the lineages closest to them based on biological function and evolutionary relationship. For example, viruses with recombinant mutations of D614G+501Y+69-70del would be considered as lineage B.1.1.7, because 501Y and 69-70del were two important mutations associated with increased transmissibility and moderately reduced neutralization in lineage B.1.1.7^1,2^. For those pseudovirus prepared on the genetic background of parental strains (e.g., Wuhan-Hu-1 from lineage B) combined with specific mutation sites (614G) in spike, such mutant was named as ‘lineage B+D614G’.

*Rationale for classifying study participants into previously-infected vaccine recipients*

Three studies reported neutralizing antibodies from serum collected from individuals who were infected with SARS-CoV-2 previously^3-5^. The majority of blood samples in Trinité’s study were collected approximately 10 months ago, long before the time when lineage B.1.1.7 firstly detected in England^6^, so we reasonably assumed that the individuals were infected with non-variants^5^. Similarly, samples from another two studies were collected in the early phase of the pandemic or before the emergence of the lineage B.1.351, therefore we assumed that those individuals were infected with parental lineages rather than mutant lineages. Thus, study participants in these three studies were defined as previously-infected vaccine recipients.

### Supplementary Tables

#### Table S1. Search strategy for three peer-reviewed databases and three preprint servers

| **Database** | **Step** | **Search strategy** | **Number of articles*** |
| --- | --- | --- | --- |
| PubMed | #1 | 2019-nCoV OR “coronavirus disease 2019” OR COVID-19 OR “severe acute respiratory syndrome coronavirus 2” OR SARS-CoV-2 | 117,925 |
|  | #2 | variant* OR mutant* OR mutation* OR “B.1.1.7” OR “20I/501Y.V1” OR “VOC 202012/01” OR “B.1.351” OR “20H/501Y.V2” OR “P.1” OR “20J/501Y.V3” OR “B.1.1.28” OR “501Y*” OR “N501Y” OR “E484K” OR “D614G” OR “69/70 deletion” OR “144Y deletion” OR “A570D” OR “P681H” OR “K417N*” OR “VUI-202012/01” OR “Kent” OR “B.1.427” OR “CA VUI1” OR “CAL.20C” OR “B.1.429” OR “B.1.526” OR “B.1.525” OR “P.2” OR “P.3” OR “B.1.616” | 1,292,118 |
|  | #3 | antibod* OR neutraliz* OR sera OR serum OR plasma OR “vaccin*” OR “immun*” | 4,463,402 |
|  | #4 | Language: English | 27,670,632 |
|  | #5 | 2020/9/1-2021/4/18 | 1,131,986 |
|  | #6 | #1 AND #2 AND #3 AND #4 AND #5 | 933 |
| Embase | #1 | 2019-nCoV OR “coronavirus disease 2019” OR COVID-19 OR “severe acute respiratory syndrome coronavirus 2” OR SARS-CoV-2 | 87,504 |
|  | #2 | variant* OR mutant* OR mutation* OR “B.1.1.7” OR “20I/501Y.V1” OR “VOC 202012/01” OR “B.1.351” OR “20H/501Y.V2” OR “P.1” OR “20J/501Y.V3” OR “B.1.1.28” OR “501Y*” OR “N501Y” OR “E484K” OR “D614G” OR “69/70 deletion” OR “144Y deletion” OR “A570D” OR “P681H” OR “K417N*” OR “VUI-202012/01” OR “Kent” OR “B.1.427” OR “CA VUI1” OR “CAL.20C” OR “B.1.429” OR “B.1.526” OR “B.1.525” OR “P.2” OR “P.3” OR “B.1.616” | 1,437,536 |
|  | #3 | antibod* OR neutraliz* OR sera OR serum OR plasma OR “vaccin*” OR “immun*” | 4,919,991 |
|  | #4 | Language: English | 24,635,636 |
|  | #5 | 2020/9/1-2021/4/18 | 1,215,720 |
|  | #6 | #1 AND #2 AND #3 AND #4 AND #5 | 605 |
| Web of Science | #1 | 2019-nCoV OR “coronavirus disease 2019” OR COVID-19 OR “severe acute respiratory syndrome coronavirus 2” OR SARS-CoV-2 | 110,574 |
|  | #2 | variant* OR mutant* OR mutation* OR “B.1.1.7” OR “20I/501Y.V1” OR “VOC 202012/01” OR “B.1.351” OR “20H/501Y.V2” OR “P.1” OR “20J/501Y.V3” OR “B.1.1.28” OR “501Y*” OR “N501Y” OR “E484K” OR “D614G” OR “69/70 deletion” OR “144Y deletion” OR “A570D” OR “P681H” OR “K417N*” OR “VUI-202012/01” OR “Kent” OR “B.1.427” OR “CA VUI1” OR “CAL.20C” OR “B.1.429” OR “B.1.526” OR “B.1.525” OR “P.2” OR “P.3” OR “B.1.616” | 1,790,443 |
|  | #3 | antibod* OR neutraliz* OR sera OR serum OR plasma OR “vaccin*” OR “immun*” | 5,300,201 |
|  | #4 | Language: English | - |
|  | #5 | 2020/1/1-2021/4/18 | - |
|  | #6 | #1 AND #2 AND #3 AND #4 AND #5 | 769 |
| Europe PMC | #1 | 2019-nCoV OR “coronavirus disease 2019” OR COVID-19 OR “severe acute respiratory syndrome coronavirus 2” OR SARS-CoV-2 | 132,048 |
|  | #2 | variant* OR mutant* OR mutation* OR “B.1.1.7” OR “20I/501Y.V1” OR “VOC 202012/01” OR “B.1.351” OR “20H/501Y.V2” OR “P.1” OR “20J/501Y.V3” OR “B.1.1.28” OR “501Y*” OR “N501Y” OR “E484K” OR “D614G” OR “69/70 deletion” OR “144Y deletion” OR “A570D” OR “P681H” OR “K417N*” OR “VUI-202012/01” OR “Kent” OR “B.1.427” OR “CA VUI1” OR “CAL.20C” OR “B.1.429” OR “B.1.526” OR “B.1.525” OR “P.2” OR “P.3” OR “B.1.616” | 1,681,877 |
|  | #3 | antibod* OR neutraliz* OR sera OR serum OR plasma OR “vaccin*” OR “immun*” | 2,622,676 |
|  | #4 | Type: Preprints | 268,552 |
|  | #5 | 2020/9/1-2021/4/18 | 1,038,347 |
|  | #6 | #1 AND #2 AND #3 AND #4 AND #5 | 641 |
| bioRxiv & medRxiv | #1 | COVID-19 OR SARS-CoV-2 | 17,140 |
|  | #2 | variant* OR mutant* OR mutation* | 65,516 |
|  | #3 | antibod* OR neutraliz* OR sera OR serum OR “vaccin*” | 58,788 |
|  | #4 | 2020/9/1-2021/4/18 | 37,105 |
|  | #5 | #1 AND #2 AND #3 AND #4 | 2,234 |

#### Table S2. Summary of characteristics of included studies

| **Author**  **(Year, Location)** | **Study participants** | | | | | | **Variants** | **Laboratory methods** | | | | | | **Outcome** | | |
| --- | --- | --- | --- | --- | --- | --- | --- | --- | --- | --- | --- | --- | --- | --- | --- | --- |
|  | **Participants (N)** | **Age (Years)** | **Vaccine (Manufacturer)** | **Dosage/Dose** | **Sampling time** | **Disease severity** | **Lineages/strains (Mutation profiles)** | **Serological assay** | **Type of Samples** | **Dilution factor (Dilution range)** | **Duplicates** | **Titrated virus** | **Cell line** | **Outcome variable** | **Geometric mean titer (95%CI)** | **Note** |
| Muik et al. (2021, Germany) | Phase1/2 trial participants (40) | Range:23-73 | BNT162b2 (Pfizer-BioNtech) | 2 doses 21d apart/30µg | 7/21d after second dose | - | Ref: Wuhan-Hu-1 (-)  V1: Lineage B.1.1.7 (full panel of mutations in Spike) | Pseudovirus neutralization assay (vsv vector) | Serum | 2-fold (1:20 to 1:2560) | 2 | Ref: 1.59x10^5;  V1: 1.30 x10^5 TU/mL | Vero 76 cell | pVNT50 | - | - |
| Wang et al. (2021, USA) | Phase 3 trial participants (14) | Median(range):  43(29-69) | mRNA-1273 (Moderna) | 2 doses 28d apart/100µg | Average (range): 8 (3-14) w after second dose | - | Ref: Wuhan-Hu-1 (19del, R683G)  V1: Lineage B.1.351 (N501Y)  V2: Lineage B.1.351 (K417N)  V3: Lineage B.1.351 (E484K)  V4: Lineage B.1.351 (K417N, E484K, N501Y) | Pseudovirus neutralization assay (lentiviral vector) | Plasma | 4-fold/5-fold (-) | - | - | 293T/ACE2.cl22 cell | NT50 | - | - |
|  | Phase 3 trial participants (6) |  | BNT162b2 (Pfizer-BioNtech) | 2 doses 21d apart/30µg |  |  |  |  |  |  |  |  |  |  |  |  |
|  | COVID-19 convalescent patients (45) | - | - | - | 1.3m/6.2m after infection | - | Ref: Wuhan-Hu-1 (19del, R683G)  V1: Lineage B.1.351 (K417N, E484K, N501Y) | Pseudovirus neutralization assay (lentiviral vector) | Plasma | 4-fold/5-fold (-) | - | - | 293TAce2 cell | NT50 | - | - |
| Wu et al. (2021, USA) | Phase 1 trial participants (8) | Range: 18-55 | mRNA-1273 (Moderna) | 2 doses 28d apart/100µg | 1w after second dose | - | Ref: Wuhan-Hu-1 with D614G variant (D614G)  V1: Lineage B.1.1.7 (D614G, 501Y)  V2: Lineage B.1.1.7 (D614G, H69/V70del, N501Y, P681HB)  V3: Lineage B.1.1.7 (H69/V70del, Y144del, N501Y, A570D, D614G, P681H, T716I, S982A, D1118H)  V4: Lineage B.1.351 (D614G, 417N, 484K, 501Y)  V5: Lineage B.1.351 (L18F, D80A, D215G, L242del, A243del, L244del, R246I, K417N, E484K, N501Y, D614G, A701V) | Pseudovirus neutralization assay (vsv vector) | Serum | 4-fold (-) | 3 | - | A549-hACE2-TMPRSS2 cell | ID50 | Ref: 1852  V5: 290 | Extract individual data from plot |
| Xie et al. (2021, USA) | Phase 1 trial participants (20) | Range: ≥16 | BNT162b2 (Pfizer-BioNtech) | 2 doses 21d apart/30µg | 2/4w after second dose | - | Ref: USA-WA1/2020(-)  V1: variant with N501Y mutation  V2: Lineage B.1.1.7 (H69/V70del, N501Y, D614G)  V3: Lineage B.1.351 (E484K, N501Y, D614G) | Plaque-reduction neutralization assay | Serum | 2-fold (1:40 to 1:1280) | - | - | Vero E6 cell | PRNT50 | - | - |
| Emary et al. (2021, UK) | Phase2/3 trial participants (-) | Range: ≥18 | ChAdOx1 nCoV-19 (AstraZeneca) | 2 doses 28d apart/  5×or 2.2×10^10^vp | - | - | Ref: Lineage Victoria (AUS-Victoria/01/2020) (S247R)  V1: Lineage B.1.1.7 (full panel of mutations in Spike) | Plaque-reduction neutralization assay | Serum | 2-fold (1:20 to 1:640) | - | 933 pfu/ml | Vero E6 Cell | ND50 | Ref: 517 (424, 631)  V1: 58 (44, 77) | Extract individual data from plot |
| Huang et al. (2021, China) | Clinical trial participants (12) | Range: 18-59^a^ | B8IBP-CorV (Sinopharm) | 2 doses 21d apart/4µg | 28d after second  dose | - | Ref1: HB02 strain (-)  Ref2: BJ01 strain (D614G)  V1: Lineage B.1.351 (D80A, L242del, A243del, L244del, R246I, K417N, E484K, N501Y, D614G, A701V) | CPE-based microneutralization assay | Serum | 2-fold (1:4 to the required concentration) | - | 100 CCID50 | Vero cell | NT50 | Ref1: 110.9 (76.7-160.2)  V1: 71.5 (51.1-100.1) | - |
|  |  | Range: 18-65^a^ | ZF2001 (Anhui Zhifei) | 3 doses /25µg | 14d after third dose | - |  |  |  |  |  |  |  |  | Ref1: 106 (75.0-150.1)  V1: 66.6 (51.0-86.9) |  |
| Hu et al. (2021, China) | COVID-19 convalescent patients (20 cases with 40 samples) | Median (range): 51.5 (45-65) | - | - | Median (range):25 (5-33) d or 230 (221-248) after symptom onset | - | Ref: Wuhan-Hu-1 (-)  V1: Lineage B.1.1.7 (H60/V70del, Y144del, N501Y, A570D, D614G, P681H, T716I, S982A, D1118H)  V2: Lineage B.1.351 (K417N, E484K, N501Y, D614G) | Pseudovirus neutralization assay (lentiviral vector) | Serum | - | 3 | 3.8×10^4^ copies in 50μL | 293T-ACE2 cell | ID50 | Ref: 825  V1: 343  V2: 148 | Extract individual data from plot |
| Li et al. (2021, China) | COVID-19 convalescent patients (49) | Median (IQR): 51 (35-63) | - | - | 43 (37-48) d after symptom onset | Mild:  3 (6.1%); Moderate: 35 (71.4%); Severe:  10 (20.4%); Critical:  1 (2.1%) | Ref1: SYSU-IHV (D614G)  Ref2: Wuhan-Hu-1 (-)  V1: Lineage B.1.1.7 (N501Y, H69/V70del, Y144del, A570D, P681H, T716I, S982A, D1118H)  V2: Lineage B.1.351 (L18F, D80A, D215G, R246I, K417N, E484K, N501Y, D614G, A701V) | Pseudovirus neutralization assay (lentiviral vector) | Serum | - | 3 | - | hACE2-HeLa cell | NT50 | - | Extract individual data from plot |
| Tada et al. (2021, USA) | COVID-19 convalescent patients (28) | - | - | - | 0, 7, 28d | - | Ref: D614G variant (D614G)  V1: Variant with H69/V70del mutation  V2: Variant with N501Y mutation  V3: Lineage B.1.1.7 (H69/V70del, N501Y, P681H)  V4: Lineage B.1.351 (D614G, L18F, D80A, D215G, 242-244del, R246I, K417N, A701V, E484K, N501Y)  V5: Variant with E484K mutation  V6: Lineage B.1.1.298 (H69/V70del, Y453F, D614G, M1229I, I692V, S1147) | Pseudovirus neutralization assay (lentiviral vector) | Serum | 2-fold (-) | - | - | ACE2.293T cell | IC50 | - | Extract individual data from plot |
|  | Vaccinated individuals (5) | - | BNT162b2 (Pfizer-BioNtech) | 2 doses 21d apart/30µg | 7d after second dose | - | Ref: D614G variant (D614G)  V1: Variant with S982A mutation  V2: Variant with N501Y mutation  V3: Lineage B.1.1.7 (H69/V70del, N501Y, P681H)  V4: Lineage B.1.351 (D614G, L18F, D80A, D215G, 242-244del, R246I, K417N, A701V, E484K, N501Y)  V5: Variant with E484K mutation | Pseudovirus neutralization assay (lentiviral vector) | Serum | 2-fold (-) | - | - | ACE2.293T cell | IC50 | - | Extract individual data from plot |
| Shen et al. (2021, USA) | COVID-19 convalescent patients (15) | - | - | - | 1-8w post resolution of COVID-19/  2-10w post most recent positive test | - | Ref: Wuhan-Hu-1 with D614G variant (D614G)  V1: Lineage B.1.1.7 (D614G, H69/V70del)  V2: Lineage B.1.1.7 (D614G, H69/V70del, N501Y)  V1: Lineage B.1.1.7 (H69/V70del, Y144del, N501Y, A570D, P681H, T716I, S982A, D1118H) | Pseudovirus neutralization assay (lentiviral vector) | Serum | 5-fold (-) | 2 | - | HEK 293T/17 cell | ID50 | Ref: 1105  V1: 1244  V2: 923  V3: 325 | - |
|  | Phase 1 trial participants (51) | Range: 18-55^a^ | mRNA-1273 (Moderna) | 2 doses 28d apart/100µg | 28d after first dose/second dose | - | Ref: Wuhan-Hu-1 with D614G variant (D614G)  V1: Lineage B.1.1.7 (D614G, H69/V70del)  V2: Lineage B.1.1.7 (D614G, H69/V70del, N501Y)  V3: Lineage B.1.1.7 (H69/V70del, Y144del, N501Y, A570D, P681H, T716I, S982A, D1118H) | Pseudovirus neutralization assay (lentiviral vector) | Serum | 5-fold (-) | 2 | - | HEK 293T/17 cell | ID50 | Ref: 1374  V1: 2896  V2: 1804  V3: 687 | - |
|  | Phase 1 trial participants (28) | Range: 18-59^a^ | NVX-CoV2373 (Novavax, Inc) | 2 doses/5μg of protein and 50μg of Matrix-M adjuvant | 2w after second dose | - | Ref: Wuhan-Hu-1 with D614G variant (D614G)  V1: Lineage B.1.1.7 (H69/V70del, Y144del, N501Y, A570D, P681H, T716I, S982A, D1118H) | Pseudovirus neutralization assay (lentiviral vector) | Serum | 5-fold (-) | 2 | - | HEK 293T/17 cell | ID50 | Ref: 975  V1: 448 | - |
| Wibmer et al. (2021, South Africa) | COVID-19 convalescent patients (44) | Range: ≥18 | - | - | - | Mild-to-moderate: 30; severe: 14 | Ref: Wuhan-Hu-1 with D614G variant (D614G)  V1: Lineage B.1.351 (K417N, E484K, N501Y, D614G)  V2: Lineage B.1.351 (L18F, D80A, D215G, 242-244del, K417N, E484K, N501Y, D614G, A701V) | Pseudovirus neutralization assay (lentiviral vector) | Plasma/  Serum | - | - | - | 293T/ACE2.MF cell | ID50 | - | - |
| Stamatatos et al. (2021, USA) | Previously infected donors (PIDs) prior to immunizations (15) | Median (IQR): 49 (45-56) | - | - | Median (IQR): 220 (125-269) d post symptom onset | Moderate: 53% (8/15), severe: 27% (4/15) | Ref: Wuhan-Hu-1 (-)  V1: Lineage B.1.351 (D80A, D215G, K417N, E484K, N501Y, D614G, A701V)  V2: Lineage B.1.351 (242-243del, D80A, D215G, K417N, E484K, N501Y, D614G, A701V) | Pseudovirus neutralization assay (lentiviral vector) | Serum | - | - | - | HEK-293T-hACE2 cell | ID50 | - | Extract individual data from plot |
|  | PIDs following immunization (15) | Median (IQR): 49 (45-56) | mRNA-1273 (Moderna) | 2 doses 28d apart/100µg | Median (IQR): 16 (14-18) d after first dose; 13 (10-17) d after second dose |  | Ref: Wuhan-Hu-1 (-)  V1: Lineage B.1.351 (D80A, D215G, K417N, E484K, N501Y, D614G, A701V)  V2: Lineage B.1.351 (242-243del, D80A, D215G, K417N, E484K, N501Y, D614G, A701V) | Pseudovirus neutralization assay (lentiviral vector) | Serum | - | - | - | HEK-293T-hACE2 cell | ID50 | - | Extract individual data from plot |
|  |  |  | BNT162b2 (Pfizer-BioNtech) | 2 doses 21d apart/30µg |  |  |  |  |  |  |  |  |  |  |  |  |
|  | Uninfected donors after immunization (13) | Median (IQR): 45 (32-51) | mRNA-1273 (Moderna) | 2 doses 28d apart/100µg | 6-28 d after second dose | - | Ref: Wuhan-Hu-1 (-)  V1: Lineage B.1.351 (D80A, D215G, K417N, E484K, N501Y, D614G, A701V)  V2: Lineage B.1.351 (242-243del, D80A, D215G, K417N, E484K, N501Y, D614G, A701V) | Pseudovirus neutralization assay (lentiviral vector) | Serum | - | - | - | HEK-293T-hACE2 cell | ID50 | - | Extract individual data from plot |
|  |  |  | BNT162b2 (Pfizer-BioNtech) | 2 doses 21d apart/30µg |  |  |  |  |  |  |  |  |  |  |  |  |
| Collier et al. (2021, UK) | Vaccinated participants (48) | Median (IQR):  63.5 tuode(47-84) | BNT162b2 (Pfizer-BioNtech) | 2 doses 21d apart/30µg | 3w after first dose | - | Ref: Wuhan-Hu-1 with D614G variant (D614G)  V1: Lineage B.1.1.7 (H69/V70del, 144del, N501Y, A570D, P681H, T716I, S982A, D1118H)  V2: Lineage B.1.1.7 with E484K mutation (E484K, H69/V70del, 144del, N501Y, A570D, P681H, T716I, S982A, D1118H)  V3: Lineage B.1.1.7 (H69/V70del, N501Y, A570D) | Pseudovirus neutralization assay (lentiviral vector) | Serum | - | 2 | - | 293T ACE2/TMPRSS2 cell | NT50 | Ref: | Extract individual data from plot |
|  | Recovered individuals (27) | - | - | - | - | - |  | Pseudovirus neutralization assay (lentiviral vector) | Serum | - | 2 | - | 293T ACE2/TMPRSS2 cell | NT50 | - | Extract individual data from plot |
| Chen et al. (2021, USA) | Convalescent adults (29) | - | - | - | 1m after infection | Mild | Ref: USA-WA1/2020 with D614G mutation (D614G)  V1: Lineage B.1.1.7 (H69/V70del, 144/145del, N501Y, A570D, D614G, P681H)  V2: Lineage B.1.351 (K417N, D614G)  V3: Lineage B.1.351 (E484K, N501Y, D614G)  V4: Lineage B.1.351 (K417N, E484K, N501Y, D614G)  V5: Lineage B.1.351 (D80A, 242/244del, R246I, K417N, E484K, N501Y, D614G, A701V)  V6: Lineage B.1.1.248 (L18F, T20N, P26S, D138Y, R190S, K417T, E484K, N501Y, D614G, H655Y, T1027I, V1176F) | Pseudovirus neutralization assay (vsv vector) | Serum | - | 2 | - | Vero-hACE2 TMPRSS2 cell | EC50 | Ref: 174  V1: 129  V2: 261  V3: 34  V4: 49  V5: 38  V6: 108 | Extract individual data from plot |
|  | Vaccinated individuals (24) | - | BNT162b2 (Pfizer-BioNtech) | 2 doses 21d apart/30µg | 1w after boosting with vaccine | - | Ref: USA-WA1/2020 with D614G mutation (D614G)  V1: Lineage B.1.1.7 (H69/V70del, 144/145del, N501Y, A570D, D614G, P681H)  V2: Lineage B.1.351 (K417N, D614G)  V3: Lineage B.1.351 (E484K, N501Y, D614G)  V5: Lineage B.1.351 (D80A, 242/244del, R246I, K417N, E484K, N501Y, D614G, A701V)  V6: Lineage B.1.1.248 (L18F, T20N, P26S, D138Y, R190S, K417T, E484K, N501Y, D614G, H655Y, T1027I, V1176F) | Pseudovirus neutralization assay (vsv vector) | Serum | - | 2 | - | Vero-hACE2 TMPRSS2 cell | EC50 | Ref: 551  V1: 272  V2: 555  V3: 132  V5: 54  V6: 233 | Extract individual data from plot |
| Garcia-Beltran et al. (2021, USA) | Vaccine recipients with 2 full doses (30) | Range: 26-66 | BNT162b2 (Pfizer-BioNtech) | 2 doses 21d apart/30µg | ≥7d after second dose | - | Ref: Wuhan-Hu-1 (-)  Ref1: Wuhan-Hu-1 with D614G mutation (D614G)  V1: Lineage B.1.1.7 (H69/V70del, Y145del, N501Y, A570D, D614G, P681H, T716I, S982A, D1118H)  V2: Lineage B.1.1.298 (H69/V70del, Y453F, D614G, M1229I, I692V)  V3: Lineage B.1.429 (S13I, W152C, L452R, D614G)  V4: Lineage P.2 (E484K, D614G, V1176F)  V5: Lineage P.1 (L18F, T20N, P26S, D138Y, R190S, K417T, E484K, N501Y, D614G, H655Y, T1027I, V1176F)  V6: Lineage B.1.351 V1 (D80A, D215G, L242-L244del, K417T, E484K, N501Y, D614G, A701V)  V7: Lineage B.1.351 V2 (L18F, D80A, D215G, L242-L244del, K417T, E484K, N501Y, D614G, A701V)  V8: Lineage B.1.351 V3 (D80A, L242-L244del, R246I, K417T, E484K, N501Y, D614G, A701V) | Pseudovirus neutralization assay (lentiviral vector) | Serum | 3-fold (1:12 to 1:8748) | - | - | 293T-ACE2 cell | NT50 | Ref: 2016 | Extract individual data from plot |
|  | Vaccine recipients with 2 full doses (35) | Range: 22-73 | mRNA-1273 (Moderna) | 2 doses 28d apart/100µg |  |  |  |  |  |  |  |  |  |  | Ref: 762 |  |
|  | Vaccine recipients with 1 dose or 2 doses within 7d (14) | Range: 23-60 | BNT162b2 (Pfizer-BioNtech) | 2 doses 21d apart/30µg | Range: 4-32 days after one dose or <7d after second dose | - |  | Pseudovirus neutralization assay (lentiviral vector) | Serum | 3-fold (1:12 to 1:8748) | - | - | 293T-ACE2 cell | NT50 | Ref: 167 | Extract individual data from plot |
|  | Vaccine recipients with 1 dose or 2 doses within 7d (27) | Range: 23-65 | mRNA-1273 (Moderna) | 2 doses 28d apart/100µg |  |  |  |  |  |  |  |  |  |  | Ref: 208 |  |
|  | Vaccine recipients with 2 full doses (24) | Range: 26-66 | BNT162b2 (Pfizer-BioNtech) | 2 doses 21d apart/30µg | ≥7d after second dose | - | Ref: Wuhan-Hu-1 (-)  V1: Lineage B.1.351 (K417N, E484K, and N401Y) | Pseudovirus neutralization assay (lentiviral vector) | Serum | 3-fold (1:12 to 1:8748) | - | - | 293T-ACE2 cell | NT50 | Ref: -  V1: 88 |  |
| Souza et al. (2021, Brazil) | COVID-19-convalescent blood-donors (19) | Median (Range): 34 (18-58) | - | - | 65d after symptom onset | - | Ref: Lineage B  (SARS-CoV-2/human/BRA/SP02/2020) (-)  V1: Lineage P.1 /12(-)  V2: Lineage P.1/30 (-) | CPE-based microneutralization assay | Plasma | 2-fold (1:20 to 1:2560) | 2 | 1000 PFU/mL | Vero cell | VNT50 | Ref: 240  V1: 40  V2: 35 | - |
|  | Phase 3 trial vaccinated participants (8) | Mean (Range): 35 (29-59) | CoronaVac  (Sinovac Biotech) | 2 doses 14d apart/3µg | 153/158d after second dose | - | Ref: Lineage B (SARS-CoV-2/human/BRA/SP02/2020) (-)  V1: Lineage P.1/12 (-)  V2: Lineage P.1/30 (-) | CPE-based microneutralization assay | Plasma | 2-fold (1:20 to 1:2560) | 2 | 1000 PFU/mL | Vero cell | VNT50 | Ref: 25  V1: <20  V2: <20 | - |
| Wang et al. (2021, USA) | COVID-19 convalescent patients (20) | Mean (range): 54 (34-79) | - | - | 1m after recovery or later | - | Ref1: USA-WA1/2020 (-) (In microneutralization assay)  Ref2: USA-WA1/2020 D614G (D614G) (In pseudovirus assay)  V1: Lineage B.1.1.7 (69-70del, 144del, N501Y, A570D, P681H, T716I, S982A, D1118H)  V2: Lineage B.1.351 (L18F, D80A, 242-244del, D215G, R246I, E484K, N501Y, K417N, A701V) | CPE-based microneutralization assay | Plasma | 5-fold (1:100 to 1:1000000) | - | - | Vero E6 cell | IC50 | - | Extract individual data from plot |
|  |  |  |  |  |  |  |  | Pseudovirus neutralization assay (vsv vector) | Plasma | - | - | - | Vero E6 cell | IC50 | - |  |
|  | Phase 1 trial participants (12) | Range: 18-55^a^ | mRNA-1273 (Moderna) | 2 doses 28d apart/100µg | 15d after second dose | - |  | CPE-based microneutralization assay | Serum | 5-fold (1:100 to 1:1000000) | - | - | Vero E6 cell | IC50 | - |  |
|  |  |  |  |  |  |  |  | Pseudovirus neutralization assay (vsv vector) | Serum | - | - | - | Vero E6 cell | IC50 | - |  |
|  | Clinical individuals (10) | - | BNT162b2 (Pfizer-BioNtech) | 2 doses 21d apart/30µg | ≥7d after second dose | - |  | CPE-based microneutralization assay | Serum | 5-fold (1:100 to 1:1000000) | - | - | Vero E6 cell | IC50 | - |  |
|  |  |  |  |  |  |  |  | Pseudovirus neutralization assay (vsv vector) | Serum | - | - | - | Vero E6 cell | IC50 | - |  |
| Cele et al. (2021, South Africa) | Participants from the first South African infection wave (14) | Range: ≥30 | - | - | range: 26-38 days post-symptom onset | - | Ref: Lineage B.1.1.117 (D614G, A688V)  V1: Lineage B.1.351 (L18F, D80A, D215G, K417N, E484K, N501Y, D614G, A701V, 242-244del) | focus reduction neutralization test assay | Plasma | 2-fold (1:100 to 1:1600) | - | 50 focus-forming units (FFU) per microwell | Vero E6 cell | PRNT50 | Ref: 344.0 (275.4-458.0)  V1: 41.1 (32.7-55.50) | - |
|  | Participants from the second South African infection wave (6) | Range: 30-69 | - | - | range: 29-43 days post-symptom onset | - | Ref: Lineage B.1.1.117 (D614G, A688V)  V1: Lineage B.1.351 (L18F, D80A, D215G, K417N, E484K, N501Y, D614G, A701V, 242-244del) | Focus reduction neutralization test assay | Plasma | 2-fold (1:100 to 1:1600) | - |  | Vero E6 cell | PRNT50 | Ref: 619.7 (517.8-771.5)  V1: 149.7 (132.1-172.8) | - |
| Edara et al. (2021, -) | Acutely infected COVID-19 patients (19) | - | - | - | 5-19d post-symptom onset | - | Ref: Lineage B.1(USA/GA-EHC-083E/2020) (D614G)  V1: Lineage B.1.351 (L18F, D80A, D215G, 242-244del, K417N, E484K, N501Y, D614G, Q677H, R682W) | Focus reduction neutralization test assay | Serum | 3-fold (1:10-) | 2 | 100-200 foci per well | VeroE6 cell | FRNT50 | Ref: 135 (<20–836)  V1: 40 (<20–433) | - |
|  | COVID-19 convalescent patients (30) | - | - | - | 1-3m post-symptom onset | - | Ref: Lineage B.1(USA/GA-EHC-083E/2020) (D614G)  V1: Lineage B.1.351 (L18F, D80A, D215G, 242-244del, K417N, E484K, N501Y, D614G, Q677H, R682W) | Focus reduction neutralization test assay | Serum | 3-fold (1:10-) | 2 | 100-200 foci per well | VeroE6 cell | FRNT50 | Ref: 288 (29–2117)  V1: 59 (<20–2363) | - |
|  | COVID-19 convalescent patients (30) | - | - | - | 3-8m post-symptom onset | - | Ref: Lineage B.1(USA/GA-EHC-083E/2020) (D614G)  V1: Lineage B.1.351 (L18F, D80A, D215G, 242-244del, K417N, E484K, N501Y, D614G, Q677H, R682W) | Focus reduction neutralization test assay | Serum | 3-fold (1:10-) | 2 | 100-200 foci per well | VeroE6 cell | FRNT50 | Ref: 107 (<20–836)  V1: 50 (<20–627) | - |
|  | Phase1 trial participants (19) | Range: >56 | mRNA-1273 (Moderna) | 2 doses 28d apart/100µg | 14d after second dose | - | Ref: Lineage B.1(USA/GA-EHC-083E/2020) (D614G)  V1: Lineage B.1.351 (L18F, D80A, D215G, 242-244del, K417N, E484K, N501Y, D614G, Q677H, R682W) | Focus reduction neutralization test assay | Serum | 3-fold (1:10-) | 2 | 100-200 foci per well | VeroE6 cell | FRNT50 | Ref: 734 (256–2868)  V1: 191 (61–830) | - |
| Planas et al. (2021, France) | Infected individuals from the Orléans Cohort (28) | Mean (Range): 56.9 (46-69) | - | - | Median (range): 89 (77-105) d post onset of symptom | Asymptomatic: 1; Mild-Moderate: 11; Severe: 13; Critical: 3 | Ref: Lineage B.1 (hCoV-19/France/GE1973/2020) (D614G)  V1: Lineage B.1.1.7 (69-70del, Y144del, N501Y, A570D, D614G, P681H, T716I, S982A, D1118H)  V2: Lineage B.1.351 (L18F, D80A, D215G, 242-244del, K417N, E484K, N501Y, D614G, A701V) | Neutralization assay | Serum | - | - | - | S-Fuse cell | ED50 | - | Extract individual data from plot |
|  |  |  |  |  | Median(range): 179 (168-197) d post onset of symptoms |  |  |  |  |  |  |  |  |  |  |  |
|  | RT-qPCR-confirmed staff from the Strasbourg Cohort (30) | Mean (range): 44.8 (25-54) | - | - | Median(range):  233 (206-258) d post symptom onset | Mild-Moderate: 30 |  |  |  |  |  |  |  |  |  |  |
|  | Vaccinated health care workers (15) | Mean (range): 55 (46-63) | BNT162b2 (Pfizer-BioNtech) | 2 doses 21d apart/30µg | 2w after first dose | - | Ref: Lineage B.1 (hCoV-19/France/GE1973/2020) (D614G)  V1: Lineage B.1.1.7 (69-70del, Y144del, N501Y, A570D, D614G, P681H, T716I, S982A, D1118H)  V2: Lineage B.1.351 (L18F, D80A, D215G, 242-244del, K417N, E484K, N501Y, D614G, A701V) | Neutralization assay | Serum | - | - | - | S-Fuse cell | ED50 | - | Extract individual data from plot |
|  | Vaccinated health care workers (16) |  |  |  | 3w after first dose |  |  |  |  |  |  |  |  |  |  |  |
|  | Vaccinated health care workers (10) |  |  |  | 1w after second dose |  |  |  |  |  |  |  |  |  |  |  |
|  | Vaccinated health care workers (15) |  |  |  | 3w after second dose |  |  |  |  |  |  |  |  |  |  |  |
| Wang et al. (2021, USA) | SARS-CoV-2 patients infected in the Spring of 2020 (20) | Mean (range):  54 (34-79) | - | - | Approximately 1m after recovery or later | - | Ref: USA-WA1/2020 (-)  V1: Lineage P.1 (D614G, K417T, E484K, N501Y, L18F, T20N, P26S, D138Y, R190S, H655Y) | Pseudovirus neutralization assay (vsv vector) | Plasma | - | - | - | Vero-E6 cell | ID50 | - | Extract individual data from plot |
|  | Phase 1 trial participants (12) | Range: 18-55 | mRNA-1273 (Moderna) | 2 doses 28d apart/100µg | 15d after second dose | - | Ref: USA-WA1/2020 (-)  V1: Lineage P.1 (D614G, K417T, E484K, N501Y, L18F, T20N, P26S, D138Y, R190S, H655Y) | Pseudovirus neutralization assay (vsv vector) | Serum | - | - | - | Vero-E6 cell | ID50 | - | Extract individual data from plot |
|  | Clinical individuals (10) | - | BNT162b2 (Pfizer-BioNtech) | 2 doses 21d apart/30µg | 7d or later after second dose | - | Ref: USA-WA1/2020 (-)  V1: Lineage P.1 (D614G, K417T, E484K, N501Y, L18F, T20N, P26S, D138Y, R190S, H655Y) | Pseudovirus neutralization assay (vsv vector) | Serum | - | - | - | Vero-E6 cell | ID50 | - | Extract individual data from plot |
| Virtanen et al. (2021, Finland) | COVID-19 patients from spring and summer 2020 (18) | - | - | - | - | 11 treated at home; 4 at hospital in regular ward; 3 at hospital in ICU | Ref: Finland/1/2020 (-)  V1: Lineage B (H49Y, DQTQTN 675-679) | CPE-based microneutralization assay | Serum | 2-fold (1:20 to 1:5120) | 2 | - | VE6-TMPRSS2-H10-cell/VE6-cell | - | - | - |
|  |  |  |  |  | 0-150 d post-symptom onset |  | Ref: Lineage B.1 (C1P1) (D614G)  V1: Lineage B.1.1.7 (full panel of mutations in Spike)  V2: Lineage B.1.351 (full panel of mutations in Spike) | CPE-based microneutralization assay | Serum | 2-fold (1:20 to 1:5120) | 2 | - |  | - | Ref: 241  V1: 202  V2: 32 | - |
|  |  |  |  |  | 150-300d post-symptom |  | Ref: Lineage B.1 (C1P1) (D614G)  V1: Lineage B.1.1.7 (full panel of mutations in Spike)  V2: Lineage B.1.351 (full panel of mutations in Spike) | CPE-based microneutralization assay | Serum | 2-fold (1:20 to 1:5120) | 2 | - |  | - | Ref: 49  V1: 76  V2: 17 | - |
| Moyo-Gwete et al. (2021, South Africa) | 501Y.V2-infected patients (22 sequence confirmed samples) | Range: ≥18 | - | - | Median (IQR): 7 (3, 11) days post PCR confirmed SARS-CoV-2 infection | Moderate | V1: Wuhan-Hu-1 with D614G mutation (D614G)  V2: Lineage B.1.351 (L18F, D80A, D215G, K417N, E484K, N501Y, D614G, A701V, 242-244del)  V3: Lineage B.1.351-RBD (K417N, E484K and N501Y, D614G)  V4: Lineage P.1 (L18F, T20N, P26S, D138Y, R190S, K417T, E484K, N501Y, D614G, H655Y, T1027I, V1176F) | Pseudovirus neutralization assay (lentiviral vector) | Plasma | - | 2 | - | HEK293T cell | ID50 | - | Extract individual data from plot |
| Trinité et al. (2021, Spain) | Individuals infected in March-2020 (Early samples) (16) | Median (IQR): 65 (55, 68) | - | - | Median (IQR): 48 (36-57) d after symptom onset | 11 hospitalized | Ref1: Wuhan-Hu-1 (-)  Ref2: Wuhan-Hu-1 with D614G mutation (D614G)  V2: Lineage B.1.1.7 (full panel of mutations in Spike) | Pseudovirus neutralization assay (lentiviral vector) | Plasma | 3-fold (1:60 to 1:14580) | 2 | 200 TCID50 | HEK293T/hACE2 cell | ID50 | - | Extract individual data from plot |
|  | Individuals infected in March-2020 (Late samples) (16) | Median (IQR): 56 (54, 62) | - | - | Median (IQR): 196 (186-207) d after symptom onset | 11 hospitalized | Ref1: Wuhan-Hu-1 (-)  Ref2: Wuhan-Hu-1 with D614G mutation (D614G)  V2: Lineage B.1.1.7 (full panel of mutations in Spike) | Pseudovirus neutralization assay (lentiviral vector) | Plasma | 3-fold (1:60 to 1:14580) | 2 | 200 TCID50 | HEK293T/hACE2 cell | ID50 | - | Extract individual data from plot |
|  | Individuals infected in Aug-2020 (16) | Median (IQR): 44 (37, 54) | - | - | Median (IQR): 44 (37-54) d after symptom onset | 11 hospitalized | Ref1: Wuhan-Hu-1 (-)  Ref2: Wuhan-Hu-1 with D614G mutation (D614G)  V2: Lineage B.1.1.7 (full panel of mutations in Spike) | Pseudovirus neutralization assay (lentiviral vector) | Plasma | 3-fold (1:60 to 1:14580) | 2 | 200 TCID50 | HEK293T/hACE2 cell | ID50 | - | Extract individual data from plot |
|  | B.1.1.7 infected patients (5 individuals with 13 samples) | Median (IQR): 79 (60, 91) | - | - | Median (IQR): 16 (8-20) d after symptom onset | 5 hospitalized | Ref1: Wuhan-Hu-1 (-)  Ref2: Wuhan-Hu-1 with D614G mutation (D614G)  V2: Lineage B.1.1.7 (full panel of mutations in Spike) | Pseudovirus neutralization assay (lentiviral vector) | Plasma | 3-fold (1:60 to 1:14580) | 2 | 200 TCID50 | HEK293T/hACE2 cell | ID50 | - | Extract individual data from plot |
|  | Previously infected and vaccinated individuals (16) | Median (IQR): 39 (29, 44) | BNT162b2 (Pfizer-BioNtech) | 2 doses 21d apart/30µg | Median (IQR): 13 (10-14) d after second dose | - | Ref1: Wuhan-Hu-1 (-)  Ref2: Wuhan-Hu-1 with D614G mutation (D614G)  V2: Lineage B.1.1.7 (full panel of mutations in Spike) | Pseudovirus neutralization assay (lentiviral vector) | Plasma | 3-fold (1:60 to 1:14580) | 2 | 200 TCID50 | HEK293T/hACE2 cell | ID50 | - | Extract individual data from plot |
|  | Uninfected and vaccinated individuals (16) | Median (IQR): 45 (30, 61) | BNT162b2 (Pfizer-BioNtech) | 2 doses 21d apart/30µg | Median (IQR): 9 (7-12) d after second dose | - | Ref1: Wuhan-Hu-1 (-)  Ref2: Wuhan-Hu-1 with D614G mutation (D614G)  V2: Lineage B.1.1.7 (full panel of mutations in Spike) | Pseudovirus neutralization assay (lentiviral vector) | Plasma | 3-fold (1:60 to 1:14580) | 2 | 200 TCID50 | HEK293T/hACE2 cell | ID50 | - | Extract individual data from plot |
| Faulkner et al. (2021, UK) | Acute D614G infection, mild/asymptomatic (17) | Median (range): 32 (26‐66) | - | - | Median (range): 28 (6‐35) d post symptom onset or first positive PCR | Mild/Asymptomatic | Ref: Lineage B (hCoV‐19/England/02/2020) (-)  V1: Lineage B.1.1.7 (hCoV‐19/England/204690005/2020) (D614G, 69‐70del, 144del, N501Y, A570D, P681H, T716I, S982A, D1118H)  V2: Lineage B.1.351 (D614G, L18F, D80A, D215G, 242‐244del, R246I, K417N, E484K, N501Y, A701V) | Fluorescence-based microneutralization assay | Serum/Plasma | 4-fold (1:40 to 1:2560) | 2 | - | Vero E6 cell | IC50 | - | Extract individual data from plot |
|  | Acute D614G infection, COVID‐19 patients (20) | Median (range): 61 (25‐73) | - | - | Median (range): 25 (14‐43) d post symptom onset or first positive PCR | - |  | Fluorescence-based microneutralization assay | Serum/Plasma | 4-fold (1:40 to 1:2560) | 2 | - | Vero E6 cell | IC50 | - | Extract individual data from plot |
|  | Acute B.1.1.7 infection patients (29) | Median (range): 57 (20‐99) | - | - | Median (range): 11 (4‐46) d post symptom onset or first positive PCR | 23 mild/ 6 asymptomatic | Ref: Lineage B (hCoV‐19/England/02/2020) (-)  V2: Lineage B.1.351 (D614G, L18F, D80A, D215G, 242‐244del, R246I, K417N, E484K, N501Y, A701V) | Fluorescence-based microneutralization assay | Serum/Plasma | 4-fold (1:40 to 1:2560) | 2 | - | Vero E6 cell | IC50 | - | Extract individual data from plot |
|  | Mild/asymptomatic D614G infected patients (31) | Median (range): 32 (26‐66) | - | - | 3m post symptom onset or first positive PCR | Mild/asymptomatic | Ref: Lineage B (hCoV‐19/England/02/2020) (-)  V1: Lineage B.1.1.7 (hCoV‐19/England/204690005/2020) (D614G, 69‐70del, 144del, N501Y, A570D, P681H, T716I, S982A, D1118H)  V2: Lineage B.1.351 (D614G, L18F, D80A, D215G, 242‐244del, R246I, K417N, E484K, N501Y, A701V) | Fluorescence-based microneutralization assay | Serum/Plasma | 4-fold (1:40 to 1:2560) | 2 | - | Vero E6 cell | IC50 | - | Extract individual data from plot |
|  | Vaccinated health care workers (29) | Median (IQR): 55 (49-58) | BNT162b2 (Pfizer-BioNtech) | 2 doses 21d apart/30µg | 3w after first dose | - | Ref: Lineage B.1 (hCoV-19/France/UN-FRA-WGS157/2020) (D614G)  V1: Lineage B.1.1.7 (full panel of mutations in Spike)  V2: Lineage B.1.351 (full panel of mutations in Spike) | CPE-based microneutralization assay | Serum | 2-fold (1:5 to 1:2560) | - | 2×10^3^ TCID50/ml | Vero E6 cell | NT100 | - | - |
|  |  |  |  |  | 7 d after second dose |  |  |  |  |  |  |  |  |  | Ref: 160  V1: 40  V2: 20 |  |
| Becker et al. (2021, Germany) | Vaccinated healthcare workers (9) | Median (IQR): 42 (16) | BNT162b2 (Pfizer-BioNtech) | 2 doses 21d apart/30µg | Median (range): 21 (-1-22) d after first dose | - | Ref: Lineage B.1.126 (D614G)  V1: Lineage B.1.351 (N501Y, K417N, E484K) | Fluorescence-based microneutralization assay | Serum | 2-fold (1:40 to 1:5120) | - | - | Caco-2 cell | VTN50 | - | Extract individual data from plot |
|  | Vaccinated healthcare workers (7) | Median (IQR): 42 (16) | BNT162b2 (Pfizer-BioNtech) | 2 doses 21d apart/30µg | Median (range):15 (-7-16) d after second dose | - | Ref: Lineage B.1.126 (D614G)  V1: Lineage B.1.351 (N501Y, K417N, E484K) | Fluorescence-based microneutralization assay | Serum | 2-fold (1:40 to 1:5120) | - | - | Caco-2 cell | VTN50 | - | Extract individual data from plot |
|  | Hospitalized PCR-confirmed individuals (6) | Median (IQR): 59 (19) | - | - | Median (range): 14 (2-85) d post positive PCR | Hospitalized | Ref: Lineage B.1.126 (D614G)  V1: Lineage B.1.351 (N501Y, K417N, E484K) | Fluorescence-based microneutralization assay | Serum | 2-fold (1:40 to 1:5120) | - | - | Caco-2 cell | VTN50 | - | Extract individual data from plot |
| Deng et al. (2021, USA) | Convalescent patients clinically diagnosed with COVID-19 (9) | - | - | - | 18-71d after symptom onset | - | Ref1: USA-WA1/2020 (-)  Ref2: SARS-CoV-2/human/USA/CA-UCSF-0001C/2020 clinical isolate (D614G)  V1: Lineage B.1.427 (L452R, S13I, W152C)  V2: Lineage B.1.429 (L452R, S13I, W152C) | Plaque-reduction neutralization assay | Plasma | 2-fold (1:100 to 1:3200) | - | - | Vero E6 cell | PRNT50 | - | - |
|  | Vaccine recipients receiving both doses of either BNT16b2 or mRNA-1273 (12) | - | BNT162b2 (Pfizer-BioNtech) | 2 doses 21d apart/30µg | 4-28d after second dose | - |  | Plaque-reduction neutralization assay | Plasma | 2-fold (1:100 to 1:3200) | - | - | Vero E6 cell | PRNT50 | - | - |
|  |  |  | mRNA-1273 (Moderna) | 2 doses 28d apart/100µg |  |  |  |  |  |  |  |  |  |  |  |  |
|  | Convalescent patients clinically diagnosed with COVID-19 (10) | - | - | - | 21-85d after symptom onset | - |  | CPE-based microneutralization assay | Plasma | 2-fold (1:20 to 1:2560) | - | 100 TCID50 | Vero E6 cell | TCID50 | - | - |
| Wang et al. (2021, China) | convalescent COVID-19 patients (23) | Mean (range): 56 (29-81) | - | - | Within 2m after symptom onset | 12 mild/11 severe | Ref: Wuhan-Hu-1 with D614G variant (D614G)  V1: Lineage B.1.1.7 (69-70del, 144del, N501Y, A570D, D614G, P681H, T716I, S982A, D1118H)  V2: Lineage B.1.351 (L18F, D80A, D215G, 242-244del, S305T, K417N, E484K, N501Y, D614G, A701V)  V3: Lineage P.1 (L18F, T20N, P26S, D138Y, R190S, K417T, E484K, N501Y, D614G, H655Y, T1027I, V1176F) | Pseudovirus neutralization assay (lentiviral vector) | Plasma | - (1: 60 to 1: 100000 /1: 200 to 1: 100000) | - | - | HeLa-ACE2 cell | ID50 | - | Extract individual data from plot |
| Liu et al. (2021, USA) | Phase1/3 trial participants (20) | Range: 23-69 | BNT162b2 (Pfizer-BioNtech) | 2 doses 21d apart/30µg | 2/4w after second dose | - | Ref: USA-WA1/2020 (-)  V1: Lineage B.1.1.7 (full panel of mutations in Spike)  V2: Lineage P.1 (full panel of mutations in Spike)  V3: Lineage B.1.351 (full panel of mutations in Spike)  V4: Lineage B.1.351 (242-244del, D614G)  V5: Lineage B.1.351 (B.1.351 RBD (K417N, E484K, N501Y), D614G) | Paque-reduction neutralization assay | Serum | 2-fold (1:40 to 1:1280) | 2 | 100 PFU | Vero E6 cell | PRNT50 | Ref: 532 (409-693)  V1: 663 (497-884)  V2: 437 (325-589)  V3: 194 (144-261)  V4: 485 (345-681)  V5: 331 (228-480) | - |
| Walls et al. (2021, USA) | Vaccine recipients (16) | - | BNT162b2 (Pfizer-BioNtech) | 2 doses 21d apart/30µg | 7-30d after second dose | - | Ref: Wuhan-Hu-1 with D614G mutation (D614G)  V1: Lineage B.1.351 (L18F, D80A, D215G, L242-L244 deletion, R246I, K417N, E484K, N501Y, A701V) | Pseudovirus neutralization assay (lentiviral vector) | Serum | 3-fold (-) | - | - | HEK293T cell | HIV IC50 | Ref: 600  V1: 67 | Extract individual data from plot |
| Wu et al. (2021, USA) | Phase 1 trial participants (8) | Range: 18-55 | mRNA-1273 (Moderna) | 2 doses 28d apart/100µg | 1w after second dose | - | Ref: Wuhan-Hu-1 with D614G mutation (D614G)  V1: Lineage P.1 (L18F, T20N, P26S, D138Y, R190S, K417T, E484K, N501Y, D614G, H655Y, T1027I, V1176F)  V2: Lineage B.1.427/ B.1.429-v1 (S13I, W152C, L452R, D614G)  V3: Lineage B.1.427/ B.1.429-v2 (S13I, P26S, W152C, L452R, D614G)  V4: Lineage B.1.1.7+E484K (H69/V70del, Y144del, E484K, N501Y, A570D, D614G, P681H, T716I, S982A, D1118H) | Pseudovirus neutralization assay (vsv-vector) | Serum | 4-fold (-) | 3 | - | A549-hACE2-TMPRSS2 cell | ID50 | - | Extract individual data from plot |
| Hoffmann et al. (2021, Germany) | Convalescent COVID-19 patients (10) | - | - | - | - | ICU | Ref: Wuhan-Hu-1 with D614G mutation (D614G)  V1: Lineage B.1.1.7 (full panel of mutations in Spike)  V2: Lineage B.1.351 (full panel of mutations in Spike)  V3: Lineage P.1 (full panel of mutations in Spike) | Pseudovirus neutralization assay (vsv-vector) | Plasma | - | - | - | Vero76 cell | NT50 | - | Extract individual data from plot |
|  | Individuals vaccinated with BioNTech/Pfizer vaccine BNT162b2 (15) | Range: 25-65 | BNT162b2 (Pfizer-BioNtech) | 2 doses 21d apart/30µg | 13-15 d after second dose | - | Ref: Wuhan-Hu-1 with D614G mutation (D614G)  V1: Lineage B.1.1.7 (full panel of mutations in Spike)  V2: Lineage B.1.351 (full panel of mutations in Spike)  V3: Lineage P.1 (full panel of mutations in Spike) | Pseudovirus neutralization assay (vsv-vector) | Serum | - | - | - | Vero76 cell | NT50 | - | Extract individual data from plot |
| Edara et al. (2021, USA) | Acutely infected COVID-19 patients (20) | Mean (range): 57 (26-85) | - | - | 5-19d post-symptom onset | Hospitalized (20% ICU) | Ref1: Lineage B.1(USA/GA-EHC-083E/2020) (D614G)  Ref2: USA-WA1/2020 (-)  V1: Lineage B.1.1.7 (full panel of mutations in Spike)  V2: Lineage B.1.1.7 (N501Y) | Focus reduction Neutralization test assay | Serum | 3-fold (1:10 to 1:65610) | 2 | - | VeroE6 cell | FRNT50 | Ref1: 110  Ref2: 186  V1: 116  V2: 141 | Extract individual data from plot |
|  | Convalescent COVID-19 individuals (20) | Mean: 45 | - | - | 32-94d post-symptom onset | 5 hospitalized and 15 non-hospitalized |  | Focus reduction neutralization test assay | Serum | 3-fold (1:10 to 1:65610) | 2 | - | VeroE6 cell | FRNT50 | Ref1: 91  Ref2: 168  V1: 145  V2: 145 | Extract individual data from plot |
|  | Phase 1 trial adult participants (14) | Range: 18-55 | mRNA-1273 (Moderna) | 2 doses 28d apart/100µg | 14d after second dose | - |  | Focus reduction neutralization test assay | Serum | 3-fold (1:10 to 1:65610) | 2 | - | VeroE6 cell | FRNT50 | Ref1: 804  Ref2: 1709  V1: 965  V2: 994 | Extract individual data from plot |
| Li et al. (2021, China) | Convalescent COVID-19 patients (15) | - | - | - | - |  | Ref: Wuhan-Hu-1 with D614G variant (D614G)  V1: Lineage B.1.351 (D614G, K417N)  V2: Lineage B.1.351 (D614G, E484K)  V3: Lineage B.1.351 (D614G, N501Y)  V4: Lineage B.1.351 (D614G, K417N, E484K)  V5: Lineage B.1.351 (D614G, K417N, N501Y)  V6: Lineage B.1.351 (D614G, E484K, N501Y)  V7: Lineage B.1.351 (D614G, K417N, E484K, N501Y)  V8: Lineage B.1.351 (D614G, D80A, D215G, E484K, N501Y, A701V)  V9: Lineage B.1.351 (L18F, K417N, D614G, D80A, D215G, E484K, N501Y, A701V)  V10: Lineage B.1.351 (242-244del, L18F, K417N, D614G, D80A, D215G, E484K, N501Y, A701V) | Pseudovirus neutralization assay (vsv vector) | Serum | 3-fold (1:30-) | - | 7.0×10^4^ TCID50/mL | Huh7 cell | ED50 | - | - |
|  | Convalescent COVID-19 patients (30) | - | - | - | 2, 5, 8m after symptom onset |  |  | Pseudovirus neutralization assay (vsv vector) | Serum | 3-fold (1:30-) | - | 7.0×10^4^ TCID50/mL | Huh7 cell | ED50 | - | - |
| Monin-Aldama et al. (2021, UK) | Vaccinated healthcare workers (40) | Median (range): 40.5 (22‐78) | BNT162b2 (Pfizer-BioNtech) | 2 doses 21d apart/30µg | 3,5w after first dose; 0,14d after second dose | - | Ref: Lineage B (England 2020/02/407073) (-)  V1: Lineage B.1.1.7 (full panel of mutations in Spike) | Pseudovirus neutralization assay (lentiviral vector) | Plasma | - | 2 | - | HeLa cell | ID50 | - | Extract individual data from plot |
|  | Vaccinated patients with solid tumor cancer (51) | Median (range): 73 (19‐94) | BNT162b2 (Pfizer-BioNtech) | 2 doses 21d apart/30µg | 3,5w after first dose; 0,14d after second dose | - | Ref: Lineage B (England 2020/02/407073) (-)  V1: Lineage B.1.1.7 (full panel of mutations in Spike) | Pseudovirus neutralization assay (lentiviral vector) | Plasma | - | 2 | - | HeLa cell | ID50 | - |  |
|  | Vaccinated patients with haematological cancer (12) | Median (range): 73 (19‐94) | BNT162b2 (Pfizer-BioNtech) | 2 doses 21d apart/30µg | 3,5w after first dose; 0,14d after second dose | - |  |  |  |  |  |  |  |  |  |  |
| Madhi et al. (2021, South Africa) | Phase Ib/II trial participants (13)  Phase Ib/II trial participants with confirmed SARS CoV-  2 infection between time of first vaccine dose and Day 42 (6) | Range: 18-65 | ChAdOx1 nCoV-19 (AstraZeneca) | 2 doses 28d apart /  5×10^10^ viral particles | 2w after second dose | - | Ref: Wuhan-Hu-1 with D614G mutation (D614G)  V1: Lineage B.1.351 (K417N, E484K, N501Y, D614G)  V2: Lineage B.1.351 (L18F, D80A, D215G, 242-244del, K417N, E484K, N501Y, D614G, A701V) | Pseudovirus neutralization assay (lentiviral vector) | Serum | - | - | - | 293T/ACE2.MF cell | ID50 | - | Extract individual data from plot |
|  |  |  |  |  |  |  | Ref: Lineage B.1.1.117 (D614G, A688V)  V1: Lineage B.1.351 (L18F, K417N, E484K, N501Y, 242-244 del) | Focus reduction neutralization test assay | Plasma | 2-fold (1:50 to 1:3200) | - | 2.5x10^4^ FFU/mL | Vero E6 cell | PRNT50 | Ref: 22  V1: 2 |  |
|  | Natural infected placebo recipients (6) | Range: 18-65 | - | - | - | - | Ref: Wuhan-Hu-1 with D614G mutation (D614G)  V1: Lineage B.1.351 (K417N, E484K, N501Y, D614G)  V2: Lineage B.1.351 (L18F, D80A, D215G, 242-244del, K417N, E484K, N501Y, D614G, A701V) | Pseudovirus neutralization assay (lentiviral vector) | Serum | - | - | - | 293T/ACE2.MF cell | ID50 | - | Extract individual data from plot |
|  |  |  |  |  |  |  | Ref: Lineage B.1.1.117 (D614G, A688V)  V1: Lineage B.1.351 (L18F, K417N, E484K, N501Y, 242-244 del) | Focus reduction neutralization test assay | Plasma | 2-fold (1:50 to 1:3200) | - | 2.5x10^4^ FFU/mL | Vero E6 cell | PRNT50 | Ref: 31  V1: 4 |  |
| Zhou et al. (2021, UK) | Convalescent COVID-19 patients (34) | - | - | - | 4–9w after admission to hospital | - | Ref: Lineage B (Human/AUS/VIC01/2020) (S247R)  V1: Lineage B.1.351 (D80A, D215G, L242-244del, K417N, E484K, N501Y, D614G, A701V) | Focus reduction neutralization test assay | Plasma | - | 2 | - | Vero (ATCC CCL-81) cell | FRNT50 | Ref: 475  V1: 36 | - |
|  | Vaccine recipients receiving both doses of Pfizer BNT16b2 (25) | Mean (range): 43 (25-63) | BNT162b2 (Pfizer-BioNtech) | 2 doses 21d apart/30µg | 7-17d after second dose | - | Ref: Lineage B (Human/AUS/VIC01/2020) (S247R)  V1: Lineage B.1.351 (D80A, D215G, L242-244del, K417N, E484K, N501Y, D614G, A701V) | Focus reduction neutralization test assay | Serum | - | 2 | - | Vero (ATCC CCL-81) cell | FRNT50 | Ref: 1105  V1: 146 | - |
|  | Vaccine recipients receiving both doses of the AstraZeneca (25) | - | ChAdOx1(AZ51222) (AstraZeneca) | 2 doses 28d apart/ 5× or 2.5×10^10^ viral particle | 14 or 28d after second dose | - | Ref: Lineage B (Human/AUS/VIC01/2020) (S247R)  V1: Lineage B.1.351 (D80A, D215G, L242-244del, K417N, E484K, N501Y, D614G, A701V) | Focus reduction neutralization test assay | Serum | - | 2 | - | Vero (ATCC CCL-81) cell | FRNT50 | Ref: 306  V1: 34 | - |
|  | Volunteers recently infected with B.1.1.7 (15) | - | - | - | 1-34d after infection | - | Ref: Lineage B (Human/AUS/VIC01/2020) (S247R)  V1: Lineage B.1.351 (D80A, D215G, L242-244del, K417N, E484K, N501Y, D614G, A701V) | Focus reduction neutralization test assay | Plasma | - | 2 | - | Vero (ATCC CCL-81) cell | FRNT50 | Ref: 432  V1: 138 | - |
| Dejnirattisai et al. (2021, UK) | Convalescent COVID-19 patients (34) | - | - | - | 4–9w following admission to hospital | - | Ref: Lineage B (Human/AUS/VIC01/2020) (S247R)  V1: Lineage P.1 (L18F, T20N, P26S, D138Y, R190S, K417T, E464K, N501Y, D614G, H655Y, T1027I, V1176F) | Focus reduction neutralization test assay | Plasma | - | 2 | - | Vero (ATCC CCL-81) cell | FRNT50 | - | - |
|  | Vaccine recipients receiving both doses of Pfizer BNT16b2 (25) | Mean (range): 43 (25-63) | BNT162b2 (Pfizer-BioNtech) | 2 doses 21d apart/30µg | 7-17 d after second dose | - | Ref: Lineage B (Human/AUS/VIC01/2020) (S247R)  V1: Lineage P.1 (L18F, T20N, P26S, D138Y, R190S, K417T, E464K, N501Y, D614G, H655Y, T1027I, V1176F) | Focus reduction neutralization test assay | Serum | - | 2 | - | Vero (ATCC CCL-81) cell | FRNT50 | - | - |
|  | Vaccine recipients receiving both doses of the AstraZeneca (25) | - | ChAdOx1(AZ51222) (AstraZeneca) | 2 doses 28d apart/  5×10^10^ viral particles | 14 or 28d after second dose | - | Ref: Lineage B (Human/AUS/VIC01/2020) (S247R)  V1: Lineage P.1 (L18F, T20N, P26S, D138Y, R190S, K417T, E464K, N501Y, D614G, H655Y, T1027I, V1176F) | Focus reduction neutralization test assay | Serum | - | 2 | - | Vero (ATCC CCL-81) cell | FRNT50 | - | - |
|  | Volunteers recently infected with B.1.1.7 (12) | - | - | - | 1-34d after infection | - | Ref: Lineage B.1.1.7 (-)  V1: Lineage P.1 (L18F, T20N, P26S, D138Y, R190S, K417T, E464K, N501Y, D614G, H655Y, T1027I, V1176F) | Focus reduction neutralization test assay | Plasma | - | 2 | - | Vero (ATCC CCL-81) cell | FRNT50 | - | - |
| Supasa et al. (2021, UK) | Convalescent COVID-19 patients (34) | - | - | - | 4-9w after admission to hospital | - | Ref: Lineage B (Human/AUS/VIC01/2020) (S247R)  V1: Lineage B.1.1.7 (full panel of mutations in Spike) | Focus reduction neutralization test assay | Plasma | - | 2 | - | Vero (ATCC CCL-81) cell | FRNT50 | V1: 162 | Extract individual data from plot |
|  | Vaccine recipients receiving both doses of the BNT16b2 (25) | Mean (range): 43 (25-63) | BNT162b2 (Pfizer-BioNtech) | 2 doses 21d apart/30µg | 7-17d after second dose | - | Ref: Lineage B (Human/AUS/VIC01/2020) (S247R)  V1: Lineage B.1.1.7 (full panel of mutations in Spike) | Focus reduction neutralization test assay | Serum | - | 2 | - | Vero (ATCC CCL-81) cell | FRNT50 | V1: 337 |  |
|  | Vaccine recipients receiving both doses of ChAdOx1 nCoV-19 (AZD1222) (25) | - | ChAdOx1(AZ51222) (AstraZeneca) | 2 doses 28d apart / 5×10^10^ viral particles | 14 or 28d after second dose | - | Ref: Lineage B (Human/AUS/VIC01/2020) (S247R)  V1: Lineage B.1.1.7 (full panel of mutations in Spike) | Focus reduction neutralization test assay | Serum | - | 2 | - | Vero (ATCC CCL-81) cell | FRNT50 | V1: 112 (14d)/  166 (28d) |  |
|  | Volunteers recently infected with B.1.1.7 (13) | - | - | - | 1-34d after infection | - | V1: Lineage B.1.1.7 (full panel of mutations in Spike) | Focus reduction neutralization test assay | Plasma | - | 2 | - | Vero (ATCC CCL-81) cell | FRNT50 | V1: 398 |  |
| Rees-Spear et al. (2021, UK) | Mild healthcare workers (18) | Median (IQR): 81 (70-87) | - | - | - | Mild | Ref1: Wuhan-Hu-1 (-)  Ref2: Wuhan-Hu-1 with D614G mutation (D614G)  V1: Lineage B.1.1.7 (H69/V70del, D614G)  V2: Lineage B.1.1.7 (N501Y, D614G)  V3: Lineage B.1.1.7 (H69/V70del, DY144, N501Y, A570D, D614G, P681H, T716I, S982A, D1118H) | Pseudovirus neutralization assay (lentiviral vector) | Serum | - | 2 | - | HEK293T/17 cell | ID50 | - | Extract individual data from plot |
|  | Hospitalized patients (18) | Median (IQR): 34 (29-44) | - | - | - | Severe (hospitalized) |  | Pseudovirus neutralization assay (lentiviral vector) | Serum | - | 2 | - | HEK293T/17 cell | ID50 | - |  |
| Sapkal et al. (2021, India) | Phase II trial participants (38) | Range: 12-65 | BBV152 (Bharat Biotech) | 2 doses, 28 d apart/6μg or 3μg | 4w after second dose | - | Ref: Lineage B.1 (hCoV-19/India/DL-NIV-770/2020) (D614G, S697F, P323L)  V1: Lineage B.1.1.7 (hCoV-19/India/NIVP1) (full panel of mutations)  V2: Lineage B.4 (hCoV-19/India/un-NIV_P2_2020Q111/2020) (T3511, M85del, M331, R27C, L37F, L11F, V1981) | Plaque-reduction neutralization assay | Serum | 4-fold (-) | - | 50-60 plaque forming units (PFU) in 0.1 ml | Vero CCL-81 cell | Log2 PRNT50 | Ref: 7.1237  V1: 6.7625  V2: 7.204 | Extract individual data from plot |
| Tang et al. (2021, USA) | Convalescent COVID-19 patients (9) | - | - | - | - | - | Ref: Lineage A (USA-WA1/2020) (-)  V1: Lineage B.1.429 (S13I, W152C, L452R, D614G)  V2: Lineage B.1.1.7 (H69/V70del, Del145, N501Y, A570D, D614G, P681H, T716I, S982A, D1118H)  V3: Lineage P.1 (L18F, T20N, P26S, D138Y, R190S, K417T, E484K, N501Y, H655Y, T1027I, D614G, V1176F)  V4: Lineage B.1.351 (L18F, D80A, D215G, 242-244del, K417N, E484K, N501Y, D614G, A701V) | Pseudovirus neutralization assay (lentiviral vector) | Plasma | - | - | 2x 10^5^ relative light units/  50μL/well | 293T-ACE2-TMPRSS2 cell | PsVNA50 | Ref: 262.8±264.1  V1: 117.8 ± 54.0  V2: 75.5 ± 53.2  V3: 58.6 ± 53.2  V4: 21.8 ± 17.9 | - |
| Lien et al. (2021, China) | Phase 1 clinical trial participants (15) | Range: 20-49 | MVC-COV1901 (Medigen) | 2 doses/low dose | 4w after second dose | - | Ref: Wuhan-Hu-1 (-)  V1: Wuhan-Hu-1 D614G (D614G)  V2: Lineage B.1.1.7 (full panel of mutations in Spike)  V3: Lineage B.1.351 (full panel of mutations in Spike) | Pseudovirus neutralization assay (lentiviral vector) | Serum | 2-fold (-) | - | - | HEK293T cell | ID50 | Ref: 311  V4: 36 | Extract individual data from plot |
|  |  |  |  | 2 doses/mild dose |  |  |  |  |  |  |  |  |  |  | Ref: 342  V4: 53 |  |
|  |  |  |  | 2 doses/high dose |  |  |  |  |  |  |  |  |  |  | Ref: 633  V4: 94 |  |
| Zhou et al. (2021, USA) | Convalescent COVID-19 patients (6) | -- | - | - | - | - | Ref: D614G variant (D614G)  V1: Lineage B.1.526 (S477N) (D614G, L5F, T95I, D253G, A701V, S477N)  V2: Lineage B.1.526 (E484K) (D614G, L5F, T95I, D253G, A701V, E484K)  V3: - (E484K) | Pseudovirus neutralization assay (lentiviral vector) | Serum | 2-fold (-) | - | 2.5×10^7^ cps | ACE2.293T cell | IC50 | Ref: 211  V1: 229  V2: 49  V3: 64 | Extract individual data from plot |
|  | Vaccine recipients (5) | - | BNT162b2 (Pfizer-BioNtech) | 2 doses 21d apart/30µg | 28d after second dose | - | Ref: D614G variant (D614G)  V1: Lineage B.1.1.7 (full panel of mutations in Spike)  V2: Lineage B.1.526 (S477N) (D614G, L5F, T95I, D253G, A701V, S477N)  V3: Lineage B.1.351 (full panel of mutations in Spike)  V4: Lineage B.1.526 (E484K) (D614G, L5F, T95I, D253G, A701V, E484K) | Pseudovirus neutralization assay (lentiviral vector) | Serum | 2-fold (-) | - | 2.5×10^7^ cps | ACE2.293T cell | IC50 | Ref: 1961  V1: 1935  V2: 1297  V3: 533  V4: 585 | Extract individual data from plot |
|  | Vaccine recipients (3) | - | mRNA-1273 (Moderna) | 2 doses 28d apart/100µg | 28d after second dose | - |  | Pseudovirus neutralization assay (lentiviral vector) | Serum | 2-fold (-) | - | 2.5×10^7^ cps | ACE2.293T cell | IC50 | Ref: 1745  V1: 1234  V2: 1146  V3: 485  V4: 457 | Extract individual data from plot |
| Skelly et al. (2021, UK) | HCWs with asymptomatic SARS-CoV-2 (12) | - | - | - | Mean (range): 28 (24-34) d post-PCR testing | Asymptomatic | Ref: Lineage Victoria (AUS-Victoria/01/2020) (-)  V1: Lineage B.1.1.7 (H69/V70del, Y144del, N501Y, A570D, D614G, P681H, T716I, S982A, D1118H)  V2: Lineage B.1.351 (D614G, L18F, D80A, D215G, 242-244del, R246I, K417N, A701V, E484K, N501Y) | Focus reduction neutralization test assay | Serum | 4-fold (-) | 4 | 100-200 FFU | Vero CCL81 cell | NT50 | Ref: 38.5  V1: 9.3  V2: <5 | Extract individual data from plot |
|  | HCWs with mild symptomatic COVID-19 (12) | - | - | - | Mean (range): 28 (24-37) d post-symptom onset | Mild |  | Focus reduction neutralization test assay | Serum | 4-fold (-) | 4 | 100-200 FFU | Vero CCL81 cell | NT50 | Ref: 438.4  V1: 133  V2: 119 |  |
|  | Vaccinated healthcare workers (11) | - | BNT162b2 (Pfizer-BioNtech) | 2 doses 21d apart/30µg | Mean (range): 29 (18-41) d after first dose | - | Ref: Lineage Victoria (AUS-Victoria/01/2020) (-)  V1: Lineage B.1.1.7 (H69/V70del, Y144del, N501Y, A570D, D614G, P681H, T716I, S982A, D1118H)  V2: Lineage B.1.351 (D614G, L18F, D80A, D215G, 242-244del, R246I, K417N, A701V, E484K, N501Y) | Focus reduction neutralization test assay | Serum | 4-fold (-) | 4 | 100-200 FFU | Vero CCL81 cell | NT50 | - |  |
|  | Vaccinated healthcare workers (25) | - | BNT162b2 (Pfizer-BioNtech) | 2 doses 21d apart/30µg | Mean (range): 8 (7-17) d after second dose | - |  | Focus reduction neutralization test assay | Serum | 4-fold (-) | 4 | 100-200 FFU | Vero CCL81 cell | NT50 | - |  |
| Jalkanen et al. (2021, Finland) | BNT162b2-vaccinated health care workers (167) | Mean (range): 43 (20-65) | BNT162b2 (Pfizer-BioNtech) | 2 doses 21d apart/30µg | Mean (range): 20 (16-28) d after first dose | - | Ref: Lineage B.1 (D614G, R682W, 674-678del)  V1: Lineage B.1.463 (F59Y, P384L, D614G, P812S)  V2: Lineage B.1.1.7 (69-70del, N501Y, D614G, T716, S982A, D1118H, Y144del, A570D, P681H, A831V)  V3: Lineage B.1.351 (T191, 243-245del, K417N, N501Y, D614G, A701V, D80A, D215G, E484K) | CPE-based microneutralization assay | Serum | 2-fold (1:20 initial dilution) | - | 100 TCID50 | Vero E6 cell | ID50 | Ref: 24  V1: 32  V2: 24  V3: 12 | Extract individual data from plot |
|  |  |  |  |  | Mean (range): 23 (13-33) d after second dose |  |  |  |  |  |  |  |  |  | Ref: 234  V1: 275  V2: 240  V3: 48 |  |
|  | Non-hospitalized, recovered COVID-19 patients (50) | Mean (range): 43 (19-93) | - | - | 14d-6w after the positive RT-qPCR test result | Non-hospitalized | Ref: Lineage B.1 (D614G, R682W, 674-678del)  V1: Lineage B.1.463 (F59Y, P384L, D614G, P812S)  V2: Lineage B.1.1.7 (69-70del, N501Y, D614G, T716, S982A, D1118H, Y144del, A570D, P681H, A831V)  V3: Lineage B.1.351 (T191, 243-245del, K417N, N501Y, D614G, A701V, D80A, D215G, E484K) | CPE-based microneutralization assay | Serum | 2-fold (1:20 initial dilution) | - | 100 TCID50 | Vero E6 cell | ID50 | Ref: 55  V1: 86  V2: 74  V3: 16 | Extract individual data from plot |
|  | BNT162b2 vaccinated health care workers with pre-existing anti-S1 IgG antibodies (9) | Mean (range): 43 (20-65) | BNT162b2 (Pfizer-BioNtech) | 2 doses 21d apart/30µg | Mean (range): 20 (16-28) d after first dose | - | Ref: Lineage B.1 (D614G, R682W, 674-678del)  V1: Lineage B.1.463 (F59Y, P384L, D614G, P812S)  V2: Lineage B.1.1.7 (69-70del, N501Y, D614G, T716, S982A, D1118H, Y144del, A570D, P681H, A831V) | CPE-based microneutralization assay | Serum | 2-fold (1:20 initial dilution) | - | 100 TCID50 | Vero E6 cell | ID50 | Ref: 435  V1: 320  V2: 101 | Extract individual data from plot |
|  |  |  |  |  | Mean (range): 23 (13-33) d after second dose |  |  |  |  |  |  |  |  |  | Ref: 682  V1: 640  V2: 132 |  |
| Zuckerman et al. (2021, Israel) | Vaccine recipients (18) | - | BNT162b2 (Pfizer-BioNtech) | 2 doses 21d apart/30µg | After second dose | - | Ref: Lineage B.1.1.50 (non-P681H strain) (D614G)  V1: Lineage B.1.1.50 (P681H strain) (A27S, D614G, P681H)  V2: Lineage B.1.1.7 (full panel of mutations in Spike) | CPE-based microneutralization assay | Serum | - | - | 100 TCID50 | VERO-E6 cell | TCID 50 | Ref: 536  V1: 281  V2: 299 | Extract individual data from plot |
| McCallum et al. (2021, Switzerland) | Vaccine recipients receiving both doses of the Moderna mRNA-1273 (11) | - | mRNA-1273 (Moderna) | 2 doses 28d apart/100µg | Range (7-27) d after second dose | - | Ref: Wuhan-Hu-1with D614G variant (D614G)  V1: Lineage B.1.427/B.1.429 (S13I, W152C, L452R) | Pseudovirus neutralization assay (lentiviral vector) | Plasma | 3-fold (-) | 2 | - | HEK293T cell | ID50 | Ref: 573  V1: 204 | Extract individual data from plot |
|  | Vaccine recipients receiving both doses of the Pfizer BNT16b2 (14) | - | BNT162b2 (Pfizer-BioNtech) | 2 doses 21d apart/30µg | Range (7-27) d after second dose | - | Ref: Wuhan-Hu-1 with D614G variant (D614G)  V1: Lineage B.1.427/B.1.429 (S13I, W152C, L452R) | Pseudovirus neutralization assay (lentiviral vector) | Plasma | 3-fold (-) | 2 | - | HEK293T cell | ID50 | Ref: 535  V1: 128 | Extract individual data from plot |
|  |  |  |  |  |  |  |  | Pseudovirus neutralization assay (vsv vector) | Plasma | 2-fold (-) | - | - | VeroE6-TMPRSS2 cell | ID50 | Ref: 257  V1: 95 |  |
|  | Naive individuals who received two doses (13) | - | BNT162b2 (Pfizer-BioNtech) | 2 doses 21d apart/30µg | Range (14-28) d after second dose | - | Ref: Wuhan-Hu-1 with D614G variant (D614G)  V1: Lineage B.1.427/B.1.429 (S13I, W152C, L452R)  V2: Lineage B.1.1.7 (full panel of mutations in Spike)  V3: Lineage B.1.351 (full panel of mutations in Spike)  V4: Lineage P.1 (full panel of mutations in Spike) | Pseudovirus neutralization assay (vsv vector) | Plasma | 2-fold (-) | - | - | VeroE6-TMPRSS2 cell | ID50 | Ref: 681  V1: 248  V2: 545  V3: 211  V4: 389 | Extract individual data from plot |
|  | Immune individuals who received two doses (5) | - | BNT162b2 (Pfizer-BioNtech) | 2 doses 21d apart/30µg | Range (14-28) d after second dose | - |  | Pseudovirus neutralization assay (vsv vector) | Plasma | 2-fold (-) | - | - | VeroE6-TMPRSS2 cell | ID50 | Ref: 2055  V1: 661  V2: 1236  V3: 838  V4: 1498 | Extract individual data from plot |
|  | Convalescent COVID-19 patients (9) | - | - | - | Range (15-28) days after symptom onset | - |  | Pseudovirus neutralization assay (vsv vector) | Plasma | 2-fold (-) | - | - | VeroE6-TMPRSS2 cell | ID50 | Ref: 348  V1: 70  V2: 145  V3: 82  V4: 55 | Extract individual data from plot |
|  | Convalescent COVID-19 patients infected with lineage B.1.1.7 (13) | - | - | - | Approximately 1m after infection | - |  | Pseudovirus neutralization assay (vsv vector) | Plasma | 2-fold (-) | - | - | VeroE6-TMPRSS2 cell | ID50 | Ref: 84  V1: 53  V2: 265  V3: 119  V4: 86 | Extract individual data from plot |
| Kuzmina et al. (2021, Israel) | COVID19 recovered individuals (10) | - | - | - | 3-9w after symptoms onset | Sever | Ref: Wuhan-Hu-1 (-)  V1: Lineage B.1.1.7 (N501Y)  V2: Lineage B.1.351 (N501Y, K417N, E484K) | Pseudovirus neutralization assay (lentiviral vector) | Serum | 4-fold (1:2000-1:128000) | 3 | - | HEK293T-ACE2 cells | NT50 | Ref: 8700  V1: 5900  V2: 1200 | Extract individual data from plot |
|  | Vaccinated individuals (5) | - | BNT162b2 (Pfizer-BioNtech) | 2 doses 21d apart/30µg | 3w after first dose | - | Ref: Wuhan-Hu-1 (-)  V1: Lineage B.1.1.7 (N501Y)  V2: Lineage B.1.351 (N501Y, K417N, E484K) | Pseudovirus neutralization assay (lentiviral vector) | Serum | 4-fold (1:2000-1:128000) | 3 | - | HEK293T-ACE2 cells | NT50 | Ref: 51000 | Extract individual data from plot |
|  | Vaccinated individuals (10) | - | BNT162b2 (Pfizer-BioNtech) | 2 doses 21d apart/30µg | 9-11d after second dose | - | Ref: Wuhan-Hu-1 (-)  V1: Lineage B.1.1.7 (N501Y)  V2: Lineage B.1.351 (N501Y, K417N, E484K) | Pseudovirus neutralization assay (lentiviral vector) | Serum | 4-fold (1:2000-1:128000) | 3 | - | HEK293T-ACE2 cells | NT50 | Ref: 99000  V1: 97000  V2: 14000 | Extract individual data from plot |
| Ikegame et al. (2021, Argentina) | Vaccine recipients (12) | Mean (Range): 39.5(25-56) | Gam-COVID-Vac (Gamaleya National Research Center for Epidemiology and Microbiology) | 2 doses 21d apart/10^11^ viral particles for each recombinant  adenovirus | 1m after second dose | - | Ref: Wuhan-Hu-1 with D614G variant (D614G)  V1: Lineage B.1.1.7 (H69/V70del, 145del, N501Y, A570D, D614G, P681H, T716I, S982A, D1118H)  V2: Lineage B.1.351 (D80A, D215G, 242-244del, K417N, E484K, N501Y, D614G, A701V)  V3: a single spike mutation (E484K) | Pseudovirus neutralization assay (vsv vector) | Serum | - | 3 | - | 293T-ACE2-TMPRSS2 cells | IC50 | Ref: 87.1 (49.5-154)  V1: 49.4 (23.4-105)  V2: 7.9 (3.2-19.5)  V3: 17.0 (8.6-33.8) | Extract individual data from plot |
| Wang et al. (2021, China) | Convalescent COVID-19 patients (34) | Median (range): 44.0 (23.0-64.0) | - | - | Median (IQR): 158.0 (149.5-164.5) after infection | Severe: 5; Non-severe: 29 | Ref: Wuhan-Hu-1 (-)  V1: Wuhan-Hu-1 with D614G variant (D614G)  V2: Lineage B.1.1.7 (full panel of mutations in Spike)  V3: Lineage B.1.351 (full panel of mutations in Spike) | Pseudovirus neutralization assay (vsv vector) | Serum | 3-fold (1:30- ) | - | 650 TCID50 | Huh7 cells | pVNT50 | - | Extract individual data from plot |
|  | Vaccinated healthcare workers (25) | Median (range): 41.0 (25.0-59.0) | BBIBP-CorV (Sinopharm) | 2 doses 21d apart/4µg with aluminum hydroxide adjuvant | 20d after second dose | - | Ref: Wuhan-Hu-1 (-)  V1: Wuhan-Hu-1 with D614G variant (D614G)  V2: Lineage B.1.1.7 (full panel of mutations in Spike)  V3: Lineage B.1.351 (full panel of mutations in Spike) | Pseudovirus neutralization assay (vsv vector) | Serum | 3-fold (1:30- ) | - | 650 TCID50 | Huh7 cells | pVNT50 | - | Extract individual data from plot |
|  | Vaccinated healthcare workers (25) | Median(range): 38.0 (23.0-58.0) | CoronaVac (Sinovac Biotech) | 2 doses 14d apart/3µg with aluminum hydroxide adjuvant | 14d after second dose | - | Ref: Wuhan-Hu-1 (-)  V1: Wuhan-Hu-1 with D614G variant (D614G)  V2: Lineage B.1.1.7 (full panel of mutations in Spike)  V3: Lineage B.1.351 (full panel of mutations in Spike) | Pseudovirus neutralization assay (vsv vector) | Serum | 3-fold (1:30- ) | - | 650 TCID50 | Huh7 cells | pVNT50 | - | Extract individual data from plot |
| Shen et al. (2021, USA) | Convalescent COVID-19 patients (14) | - | - | - | 1-8w after resolution of infection/2-10w after the most recent positive SARS-CoV-2 test | - | Ref: D614G strain (D614G)  V1: Lineage B.1.429 (D614G, S13I, W152C, L452)  V2: Lineage B.1.351 (D614G, L18F, D80A, D215G, 242-244del, R246I, K417N, E484K, N501Y, A701V) | Pseudovirus neutralization assay (lentiviral vector) | Serum | 5-fold (-) | 2 | - | 293T cells | ID50 | - | - |
|  | Vaccine recipients (26) | Range: >18 | mRNA-1273 (Moderna) | 2 doses 28d apart / 100µg | 28d after second dose | - | Ref: D614G strain (D614G)  V1: Lineage B.1.429 (D614G, S13I, W152C, L452)  V2: Lineage B.1.351 (D614G, L18F, D80A, D215G, 242-244del, R246I, K417N, E484K, N501Y, A701V) | Pseudovirus neutralization assay (lentiviral vector) | Serum | 5-fold (-) | 2 | - | 293T cells | ID50 | - | - |
|  | Vaccine recipients (23) | - | NVX-CoV2373 (Novavax) | 2 dose / 5µg of protein and 50µg of Matrix-M adjuvant | 14d after second dose | - | Ref: D614G strain (D614G)  V1: Lineage B.1.429 (D614G, S13I, W152C, L452)  V2: Lineage B.1.351 (D614G, L18F, D80A, D215G, 242-244del, R246I, K417N, E484K, N501Y, A701V) | Pseudovirus neutralization assay (lentiviral vector) | Serum | 5-fold (-) | 2 | - | 293T cells | ID50 | - | - |
| Betton et al. (2021, France) | Patients hospitalized with COVID-19 (107) | Mean (sd): 58.7  (14.0) | - | - | 6m post infection | Hospitalized | Ref: Lineage B.1 (hCoV-19/France/GE1973/2020) (D614G)  V1: Lineage B.1.1.7 (full panel of mutations in Spike)  V2: Lineage B.1.351 (full panel of mutations in Spike)  V3: Lineage P.1 (full panel of mutations in Spike) | Fluorescence-based micrneutralisation assay | Serum | - (1:30- ) | - | - | S-Fuse cells | ID50 | - | Extract individual data from plot |
| Bates et al. (2021, USA) | Vaccine recipients receiving both doses (51) | Median (range): 50 (21-82) | BNT162b2 (Pfizer-BioNtech) | 2 doses 21d apart / 30µg | 14-15d after second dose | - | Ref: USA-WA1/2020 (-)  V1: Lineage B.1.1.7 (N501Y)  V2: Lineage B.1.351 (K417N, E484K, N501Y) | Focus reduction neutralization test assay | Serum | 4-fold (1:10 -1:2560) | 2 | 50 FFU | Vero E6 cells | FRNT50 | - | Extract individual data from plot |
|  | Convalescent patients clinically diagnosed with COVID-19 (54) | Median (range): 56 (1-88) | - | - | Median (range): 188.5 (1-302) d after positive PCR test | 17 hospitalized; 37 non-hospitalized | Ref: USA-WA1/2020 (-)  V1: Lineage B.1.1.7 (N501Y)  V2: Lineage B.1.351 (K417N, E484K, N501Y) | Focus reduction neutralization test assay | Serum | 4-fold (1:10 -1:2560) | 2 | 50 FFU | Vero E6 cells | FRNT50 | - | Extract individual data from plot |
| Stankov et al. (2021, Germany) | Patients with severe COVID-19 (3) | Range: 19-84 | - | - | >3w since diagnosis | - | Ref: Wuhan-Hu-1 with D614G mutation (D614G)  V1: Lineage B.1.1.7 (full panel of mutations in Spike)  V2: Lineage B.1.351 (full panel of mutations in Spike)  V3: Lineage P.1 (full panel of mutations in Spike) | Pseudovirus neutralization assay (vsv vector) | Plasma | - | - | - | Vero cells | NT50 | - | Extract individual data from plot |
|  | Vaccine recipients receiving both doses (19) | Mean(range): 40.6 (22-66) | BNT162b2 (Pfizer-BioNtech) | 2 doses 21d apart / 30µg | Mean (range): 17.6 (2-27) d after first dose; or 21 (6 -36) d after second dose | - | Ref: Wuhan-Hu-1 with D614G mutation (D614G)  V1: Lineage B.1.1.7 (full panel of mutations in Spike)  V2: Lineage B.1.351 (full panel of mutations in Spike)  V3: Lineage P.1 (full panel of mutations in Spike) | Pseudovirus neutralization assay (vsv vector) | Plasma | - | - | - | Vero cells | NT50 | - | Extract individual data from plot |
| Goel et al. (2021, USA) | SARS-CoV-2 naive individuals (33) | Mean (range): 37.3 (23-67) | BNT162b2 (Pfizer-BioNtech) | 2 doses 21d apart / 30µg | 15d after first dose / 0d after second dose / 7d after second dose | - | Ref: D614G variant (D614G)  V1: Lineage B.1.351 (full panel of mutations in Spike) | Pseudovirus neutralization assay (vsv vector) | Serum | 2-fold (-) | ≥2 | 100-300 focus forming units/well | Vero E6 cells | FRNT50 | - | - |
|  |  |  | mRNA-1273 (Moderna) | 2 doses 28d apart / 100µg |  |  |  |  |  |  |  |  |  |  |  |  |
|  | SARS-CoV-2 recovered individuals (11) | Mean (range): 34.7 (23-58) | BNT162b2 (Pfizer-BioNtech) | 2 doses 21d apart / 30µg | Range (65-275) d after infection / 15d after first dose / 0d after second dose / 7d after second dose | - | Ref: D614G variant (D614G)  V1: Lineage B.1.351 (full panel of mutations in Spike) | Pseudovirus neutralization assay (vsv vector) | Serum | 2-fold (-) | ≥2 | 100-300 focus forming units/well | Vero E6 cells | FRNT50 | - | - |
|  |  |  | mRNA-1273 (Moderna) | 2 doses 28d apart / 100µg |  |  |  |  |  |  |  |  |  |  |  |  |

Abbreviations: vesicular stomatitis virus: vsv vector; m: months; w: weeks; d: days

#### Table S3. Definition of five groups of study participants

| **Study participants** | **Definition** |
| --- | --- |
| Non-variant-infected individuals | A group of people who have been infected with SARS-CoV-2 parental strains, including acutely-infected/convalescent COVID-19 patients and asymptomatic cases confirmed by either virology or serology. |
| Variant-infected individuals | A group of people who have been infected with variant strains confirmed by either virology or serology, such as B.1.1.7/B.1.351-infected COVID-19 patients. |
| Uninfected vaccine recipients | A group of people who have been administrated with vaccines but have not been infected with SARS-CoV-2. |
| Previously-infected vaccine recipients | A group of people who have been administrated vaccines and have been infected with SARS-CoV-2 parental strains confirmed by either virology or serology. |

#### Table S4. Summary of SARS-CoV-2 lineages involved in included studies

| **PANGO Lineages** | **Virus name (GISAID/Genebank ID)** | **Country (Location)** | **Major mutations** | **Ref** |
| --- | --- | --- | --- | --- |
| Lineage A.1 | hCoV-19/USA/WA1/2020  (EPI_ISL_404895) | USA (Washington) | - | Xie et al.^7^, Wang et al.^8^, Wang et al.^9^, Deng et al.^10^, Liu et al.^11^, Edara et al.^12^, Tang et al.^13^, Bates et al.^14^ |
| Lineage A.1+D614G | - | - | D614G | Chen et al.^15^, Wang et al.^8^ |
| Lineage B | hCoV-19/Wuhan/IVDC-HB-01/2019 (EPI_ISL_402119)  hCoV-19/Australia/VIC01/2020 (EPI_ISL_406844)  hCoV-19/Brazil/SP-02/2020 (EPI_ISL_413016) | China (Hubei, Wuhan)  Australia (Victoria)  Brazil (São Paulo) | - | Muik et al.^2^, Wang et al.^16^, Emary et al.^17^, Huang et al.^18^, Hu et al.^19^, Li et al.^20^, Stamatatos et al.^4^, Garcia-Beltran et al.^21^, Souza et al.^22^, Virtanen et al.^23^, Trinité et al.^5^, Faulkner et al.^24^, Monin-Aldama et al.^25^, Zhou et al.^26^, Rees-Spear et al.^27^, Lien et al.^28^, Skelly et al.^29^, Kuzmina et al.^30^, Wang et al.^31^ |
| Lineage B+D614G | - | - | D614G | Wu et al.^32^, Tada et al.^33^, Shen et al.^34^, Wibmer et al.^35^, Collier et al.^36^, Garcia-Beltran et al.^21^, Trinité et al.^5^, Wang et al.^37^, Walls et al.^38^, Wu et al.^39^, Hoffmann et al.^40^, Li et al.^41^, Madhi et al.^3^, Rees-Spear et al.^27^, Lien et al.^28^, Zhou et al.^42^, McCallum et al.^43^, Ikegame et al.^44^, Moyo-Gwete et al.^45^, Wang et al.^31^, Shen et al.^46^, Stankov et al.^47^, Goel et al.^48^ |
| Lineage B.1 | hCoV-19/USA/GA-EHC-083E/2020 (EPI_ISL_454690)  hCoV-19/France/GES-1973/2020  (EPI_ISL_414631) | USA (Georgia)  France (Grand-Est) | D614G | Li et al.^20^, Edara et al.^12,49^, Planas et al.^50^, Virtanen et al.^23^, Deng et al.^10^, Edara et al.^12^, Sapkal et al.^51^, Jalkanen et al.^52^, Zuckerman et al.^53^, Betton et al.^54^ |
| Lineage B.1.1.117 ^a^ | hCoV-19/South Africa/KRISP-K002868/2020 (EPI_ISL_602622) | South Africa (KwaZulu-Natal) | D614G, A688V | Cele et al.^55^, Madhi et al.^3^ |
| Lineage B.1.1.29 ^a^ | hCoV-19/Beijing/IVDC-01-06/2020 (EPI_ISL_469254) | China (Beijing) | D614G | Huang et al.^18^ |
| Lineage B.1.1.298 (Mink cluster V) | hCoV-19/Denmark/DCGC-2934/2020 (EPI_ISL_616695) | Denmark (Nordjylland) | 69-70del, Y453F, D614G, M1229I, I692V | Tada et al.^33^, Garcia-Beltran et al.^21^ |
| Lineage B.1.1.50 ^a^ |  |  |  | Zuckerman et al.^53^ |
| Lineage B.1.1.7 | hCoV-19/England/MILK-9E05B3/2020 (EPI_ISL_601443) | United Kingdom (England) | N501Y, 69-70del, P681H, D614G | Muik et al.^2^, Wu et al.^32^, Xie et al.^7^, Emary et al.^17^, Hu et al.^19^, Li et al.^20^, Tada et al.^33^, Shen et al.^34^, Collier et al.^36^, Chen et al.^15^, Garcia-Beltran et al.^21^, Wang et al.^8^, Planas et al.^50^, Virtanen et al.^23^, Trinité et al.^5^, Faulkner et al.^24^, Wang et al.^37^, Liu et al.^41^, Hoffmann et al.^40^, Edara et al.^12^, Monin-Aldama et al.^25^, Supasa et al.^1^, Rees-Spear et al.^27^, Sapkal et al.^51^, Tang et al.^13^, Lien et al.^28^, Zhou et al.^42^, Skelly et al.^29^, Jalkanen et al.^52^, Zuckerman et al.^53^, McCallum et al.^43^, Ikegame et al.^44^, Wang et al.^31^, Betton et al.^54^, Bates et al.^14^, Stankov et al.^47^ |
| Lineage B.1.1.7+E484K | - | - | - | Wu et al.^39^ |
| Lineage B.1.126 ^a^ | hCoV-19/Germany/NI-IOV-P5839/2020 (EPI_ISL_883187) | Europe (Germany) | D614G | Becker et al.^56^ |
| Lineage B.1.351 | hCoV-19/South Africa/NHLS-UCT-GS-1067/2020 (EPI_ISL_700428) | South Africa (Western Cape) | N501Y, E484K, K417N, D614G | Wang et al.^16^, Wu et al.^32^, Xie et al.^7^, Huang et al.^18^, Hu et al.^19^, Li et al.^20^, Tada et al.^33^, Wibmer et al.^35^, Stamatatos et al.^4^, Chen et al.^15^, Garcia-Beltran et al.^21^, Wang et al.^8^, Cele et al.^55^, Edara et al.^49,57^, Planas et al.^50^, Virtanen et al.^23^, Faulkner et al.^24^, Becker et al.^56^, Wang et al.^37^, Liu et al.^11^, Walls et al.^38^, Hoffmann et al.^40^, Li et al.^41^, Madhi et al.^3^, Zhou et al.^26^, Tang et al.^13^, Lien et al.^28^, Zhou et al.^42^, Skelly et al.^29^, Jalkanen et al.^52^, McCallum et al.^43^, Kuzmina et al.^30^, Ikegame et al.^44^, Moyo-Gwete et al.^45^, Wang et al.^31^, Shen et al.^46^, Betton et al.^54^, Bates et al.^14^, Stankov et al.^47^, Goel et al.^48^ |
| Lineage B.1.427/B.1.429 | - | - | L452R, S13I, W152C | Garcia-Beltran et al.^21^, Deng et al.^10^, Wu et al.^39^, Tang et al.^13^, McCallum et al.^43^, Shen et al.^46^ |
| Lineage B.1.463 | SARS-CoV-2/human/FIN/SR121-VE6-H10/2020 (GenBank: MW717676) | Finland | F59Y, P384L, D614G, P812S | Jalkanen et al.^52^ |
| Lineage B.1.526 |  |  | D614G, L5F, T95I, D253G, A701V, E484K (or S477N) | Zhou et al.^42^ |
| Lineage B.4 | hCoV-19/India/un-NIV_P2_2020Q111/2020 (EPI_ISL_1061434) | India | - | Sapkal et al.^51^ |
| Lineage P.1 (Lineage B.1.1.28 P1) | hCoV-19/Japan/IC-0561/2021 (EPI_ISL_792680) ^b^ | Brazil | N501Y, E484K, K417N/T, D614G | Chen et al.^15^, Garcia-Beltran et al.^21^, Souza et al.^22^, Wang et al.^9^, Wang et al.^37^, Liu et al.^11^, Wu et al.^39^, Hoffmann et al.^40^, Dejnirattisai et al.^58^, Tang et al.^13^, McCallum et al.^43^, Moyo-Gwete et al.^45^, Betton et al.^54^, Stankov et al.^47^ |
| Lineage P.2 (Lineage B.1.1.28 P2) | hCoV-19/Brazil/CE-IEC-177339/2020 (EPI_ISL_918536) | Brazil | E484K, D614G, V1176F | Garcia-Beltran et al.^21^ |

^a^ These lineages belong to the descendants of B.1 lineage and carry mild mutation sites that had minor impact on neutralization escape, therefore were classified into B.1 lineage in our analysis.

^b^ “Airport Quarantine Station in Japan. COVID-19 positive case who entered from Brazil” from GISAID.

^c^ Detected in some sequences but not all

#### Table S5. Summary of vaccines ^59^

| **No** | **Vaccine** | **Manufacturer** | **Platform** | **Antigen** | **Virus (GenBank ID)** | **Dose** | **Schedule** |
| --- | --- | --- | --- | --- | --- | --- | --- |
| 1 | mRNA-1273 | Moderna (US) | RNA | Full-length spike (S) protein with proline substitutions | Wuhan-Hu-1 (MN908947.3) | 100 μg | 2 Doses, 28 d apart |
| 2 | BNT162b2 | Pfizer-BioNTech (US) | RNA | Full-length S protein with proline substitutions | Wuhan-Hu-1 (MN908947.3) | 30 μg | 2 Doses, 21 d apart |
| 3 | ChAdOx1 nCoV-19 (AZD1222) | AstraZeneca/Oxford (UK) | Non-replicating vector | Replication-deficient chimpanzee adenoviral vector with the SARS-CoV-2 S protein | Wuhan-Hu-1 (MN908947) | 5 × 10^10 Viral particles (standard dose) | 2 Doses, 28 d apart |
| 4 | Gam-COVID-Vac (Sputnik V) | Gamaleya National Research Center for Epidemiology and Microbiology (Russia) | Non-replicating vector | Full-length SARS-CoV-2 glycoprotein S carried by adenoviral vectors | - | 10^11 Viral particles per dose for each recombinant adenovirus | 2 Doses (first, rAd26; second, rAd5), 21 d apart |
| 5 | CoronaVac | Sinovac Biotech (China) | Inactivated | Inactivated CN02 strain of SARS-CoV-2 created from Vero cells | SARS-CoV-2/human/CHN/CN2/2020 (MT407650) | 3 μg with aluminum hydroxide adjuvant | 2 Doses, 14 d apart |
| 6 | BBIBP-CorV | Sino pharm (China) | Inactivated | Inactivated HB02 strain of SARS-CoV-2 created from Vero cells | HB02 (19nCoV-CDC-Tan-HB02) | 4 μg with aluminum hydroxide adjuvant | 2 Doses, 21 d apart |
| 7 | BBV152 (COVAXIN) | Bharat Biotech (India) | Inactivated | Inactivated NIV-2020-770 strain of SARS-CoV-2 created from Vero cells (contains Asp614Gly mutation) | NIV-2020-770 | 6 μg With Algel-IMDG adjuvant | 2 Doses, 28 d apart |
| 8 | NVX-CoV2373 | Novavax, Inc (US) | Protein subunit | Recombinant full-length, prefusion S protein | Wuhan-Hu-1 (MN908947); nucleotides 21563–25384 | 5 μg of protein and 50 μg of Matrix-M adjuvant | 2 Doses, 21 d apart |
| 9 | ZF2001 | Anhui Zhifei Longcom Biopharmaceutical (China) | Protein subunit | RBD-based protein | SARS-CoV-2 surface glycoprotein (YP_009724390); RBD antigen (residues 319 - 537) | 25 μg | 3 Doses, 30 d apart |
| 10 | MVC-COV1901 | Medigen | Protein Subunit | Spike protein S-2P with CpG 1018 | Wuhan-Hu-1 (MN908947) | 50 μg S-2P+750 μg CpG 1018 + 375 μg alum | 2 Doses, 28 d apart |

#### Table S6. Variables used in univariate and multivariate analysis

| **Variables** | **Subgroup Items** | **Details** | **Whether quantitatively measure** |
| --- | --- | --- | --- |
| Uninfected vaccine recipients | | | |
| Platforms | Non-replicating vector  RNA  Inactivated  Protein subunit | ChAdOx1 nCoV-19, Gam-COVID-Vac  BNT162b2, mRNA-1273  BBIBP-CorV, CoronaVac, BBV152  ZF2001, NVX-CoV2373, MVC-COV1901 | Yes |
| Sampling interval after second dose | < 14 days  14-90 days  > 90 days | Based on a study that evaluated antibody persistence through 6 months after vaccination^60^. | Yes |
| Non-variant-infected patients | | | |
| Sampling interval post symptom onset | ≤ 30 days  31-90 days  > 90 days | Figure S9 | Yes |
| Severity | Hospitalized  Non-hospitalized | According to WHO clinical management criteria^1^, asymptomatic/mild and moderate refers to non-hospitalized, severe and critical refers to hospitalized. | Yes |

#### Table S7. Summary characteristics of included articles

| **Characteristic** | **No. of Studies (%)** | **No. of samples (%)** |
| --- | --- | --- |
| Study group ^a^ |  |  |
| Non-variant-infected individuals | 41 (73.2) | 3440 (40.0) |
| Variant-infected individuals | 8 (14.3) | 288 (3.4) |
| Uninfected vaccine recipients | 44 (78.6) | 4697 (54.7) |
| Non-replicating vector vaccine | 3 (5.4) | 199 (4.2) |
| Inactivated vaccine | 4 (7.1) | 357 (7.6) |
| mRNA vaccine | 36 (64.3) | 3800 (80.9) |
| Protein subunit vaccine | 4 (7.1) | 341 (7.3) |
| Previously-infected vaccine recipients | 6 (10.7) | 165 (1.9) |
| Neutralization assay ^a^ |  |  |
| Live virus assay | 24 (42.9) | 3676 (42.8) |
| Lentivirus-vector pseudovirus assay | 22 (39.3) | 3282 (38.2) |
| VSV-vector pseudovirus assay ^b^ | 13 (23.2) | 1632 (19.0) |
| Pango lineage ^a^ |  |  |
| Reference strains | - | 3063 (35.7) |
| A.1 | 8 (14.3) | 328 (3.8) |
| A.1+D614G | 2 (3.6) | 95 (1.1) |
| B | 19 (33.9) | 944 (11.0) |
| B.1 ^c^ | 15 (26.8) | 741 (8.6) |
| B+D614G | 23 (41.1) | 934 (10.9) |
| B.4 | 1 (1.8) | 21 (0.2) |
| Variants of concern | - | 5137 (59.8) |
| B.1.1.7 ^d^ | 37 (66.1) | 1852 (21.6) |
| B.1.351 | 40 (71.4) | 2422 (28.2) |
| P.1 | 14 (25.0) | 570 (6.6) |
| B.1.427/B.1.429 | 6 (10.7) | 293 (3.4) |
| Variants of Interest | - | 132 (1.5) |
| B.1.526 | 1 (1.8) | 28 (0.3) |
| P.2 | 1 (1.8) | 104 (1.2) |
| Other variants ^e^ | 3 (5.4) | 258 (3.0) |
| Country |  |  |
| Argentina | 1 (1.8) | 36 (0.4) |
| Brazil | 1 (1.8) | 81 (0.9) |
| China | 7 (12.5) | 1045 (12.2) |
| Finland | 2 (3.6) | 898 (10.5) |
| France | 2 (3.6) | 582 (6.8) |
| Germany | 4 (7.1) | 286 (3.3) |
| India | 1 (1.8) | 97 (1.1) |
| Israel | 2 (3.6) | 98 (1.1) |
| South Africa | 4 (7.1) | 376 (4.4) |
| Spain | 1 (1.8) | 211 (2.5) |
| Switzerland | 1 (1.8) | 278 (3.2) |
| UK | 9 (16.1) | 1080 (12.6) |
| USA | 21 (37.5) | 3522 (41.0) |
| Status of studies |  |  |
| Peer-review | 35 (62.5) | 5584 (65.0) |
| Preprint | 21 (37.5) | 3006 (35.0) |

^a^ Some studies may include multiple study groups, neutralization assays, and lineages.

^b^ VSV, vesicular stomatitis virus.

^c^ Lineage B.1 includes the descendants of B.1 lineage carrying mild mutation sites that had little impact on neutralization escape, such as B.1.1.29, B.1.1.117, B.1.126, and B.1.1.50.

^d^ Lineage B.1.1.7 includes the Lineage B.1.1.7 with the E484K mutation site here.

^e^ Includes B.1.1.298 and B.1.463 variant.

#### Table S8. Multivariate regression of neutralizing antibodies of non-variant-infected patients

|  | **e^β^ (95% CI) ^b^** | **p value** |
| --- | --- | --- |
| **Live virus neutralization test (n=574)** | | |
| Pango lineage |  |  |
| Reference strains ^a^ | Reference |  |
| B.1.1.7 | 1.169 (0.887, 1.541) | 0.269 |
| B.1.351 | 0.299 (0.225, 0.396) | <0.001 |
| P.1 | 1.042 (0.744, 1.459) | 0.812 |
| Sampling interval post symptom onset | | |
| 31-90 days | Reference |  |
| 0-30 days | 1.883 (0.945, 3.752) | 0.073 |
| >90 days | 5.563 (2.852, 10.85) | <0.001 |
| Clinical severity |  |  |
| Non-hospitalized | Reference |  |
| Hospitalized | 2.229 (1.717, 2.895) | <0.001 |
| **Lentivirus-vector pseudovirus neutralization test (n=500)** | | |
| Pango lineage |  |  |
| Reference strains ^a^ | Reference |  |
| B.1.1.7 | 0.893 (0.596, 1.339) | 0.585 |
| B.1.351 | 0.139 (0.097, 0.199) | <0.001 |
| Sampling interval post symptom onset | | |
| 31-90 days | Reference |  |
| 0-30 days | 0.376 (0.237, 0.596) | <0.001 |
| >90 days | 0.324 (0.204, 0.516) | <0.001 |
| **VSV-vector pseudovirus neutralization test (n=200)** | | |
| Pango lineage |  |  |
| Reference strains ^a^ | Reference |  |
| B.1.1.7 | 0.85 (0.489, 1.477) | 0.564 |
| B.1.351 | 0.414 (0.272, 0.631) | <0.001 |
| P.1 | 0.497 (0.269, 0.916) | <0.05 |
| Sampling interval post symptom onset | | |
| 31-90 days | Reference |  |
| 0-30 days | 0.195 (0.138, 0.276) | <0.001 |

^a^ Included lineage B, B.1, B+D614G, B.1.1.29, B.1.1.117, B.1.126, B.1.1.50, B.4;

^b^ β higher than 0 indicates that the predictors were associated with increases of SARS-CoV-2 neutralizing antibodies by (e^β^–1) folds; β lower than 0 indicates the predictors were associated with decreases of SARS-CoV-2 neutralizing antibodies by (1–e^β^) folds.

#### Table S9. Multivariate regression of neutralizing antibodies of uninfected vaccine recipients

|  | **e^β^ (95% CI) ^b^** | **p value** |
| --- | --- | --- |
| **Live virus neutralization test (n=676)** | | |
| Pango lineage |  |  |
| Reference strains ^a^ | Reference |  |
| B.1.1.7 | 1.059 (0.82, 1.369) | 0.659 |
| B.1.351 | 0.293 (0.232, 0.37) | <0.001 |
| P.1 | 0.371 (0.271, 0.51) | <0.001 |
| B.1.427/ B.1.429 | 0.411 (0.206, 0.818) | <0.05 |
| Vaccine platform |  |  |
| Non-replicating vector | Reference |  |
| Inactivated | 11.397 (6.959, 18.665) | <0.001 |
| mRNA | 61.504 (38.596, 98.007) | <0.001 |
| Protein subunit | 11.705 (6.53, 20.979) | <0.001 |
| Sampling interval after last dose | | |
| <14 days | Reference |  |
| 14-90 days | 0.905 (0.676, 1.213) | 0.505 |
| >90 days | 0.18 (0.099, 0.326) | <0.001 |
| **Lentivirus-vector pseudovirus neutralization test (n=955)** | | |
| Pango lineage |  |  |
| Reference strains ^a^ | Reference |  |
| B.1.1.7 | 0.534 (0.402, 0.711) | <0.001 |
| B.1.351 | 0.105 (0.079, 0.141) | <0.001 |
| P.1 | 0.244 (0.138, 0.431) | <0.001 |
| B.1.427/ B.1.429 | 0.382 (0.259, 0.564) | <0.001 |
| P.2 | 0.16 (0.09, 0.283) | <0.001 |
| B.1.526 | 0.404 (0.173, 0.943) | <0.05 |
| Vaccine platform |  |  |
| mRNA | Reference |  |
| Protein subunit | 0.287 (0.217, 0.378) | <0.001 |
| Sampling interval after last dose | | |
| <14 days | Reference |  |
| 14-90 days | 6.765 (5.081, 9.008) | <0.001 |
| **Lentivirus-vector pseudovirus neutralization test (n=589)** | | |
| Pango lineage |  |  |
| Reference strains ^a^ | Reference |  |
| B.1.1.7 | 0.898 (0.711, 1.135) | 0.369 |
| B.1.351 | 0.376 (0.299, 0.473) | <0.001 |
| P.1 | 0.901 (0.6, 1.353) | 0.615 |
| B.1.427/ B.1.429 | 1.489 (0.84, 2.64) | 0.174 |
| Vaccine platform |  |  |
| Non-replicating vector | Reference |  |
| mRNA | 0.883 (0.599, 1.3) | 0.529 |
| Protein subunit | 8.678 (5.717, 13.174) | <0.001 |
| Sampling interval after last dose | | |
| <14 days | Reference |  |
| 14-90 days | 0.856 (0.657, 1.115) | 0.25 |

^a^ Included lineage B, B.1, B+D614G, B.1.1.29, B.1.1.117, B.1.126, B.1.1.50, B.4;

^b^ β higher than 0 indicates that the predictors were associated with increases of SARS-CoV-2 neutralizing antibodies by (e^β^–1) folds; β lower than 0 indicates the predictors were associated with decreases of SARS-CoV-2 neutralizing antibodies by (1–e^β^) folds.

### Supplementary Figures

#### Fig S1. Summary of countries reporting variant cases, as of 13 April 2021

Data source: *COVID-19 Weekly Epidemiological Update* published by WHO ^61^.

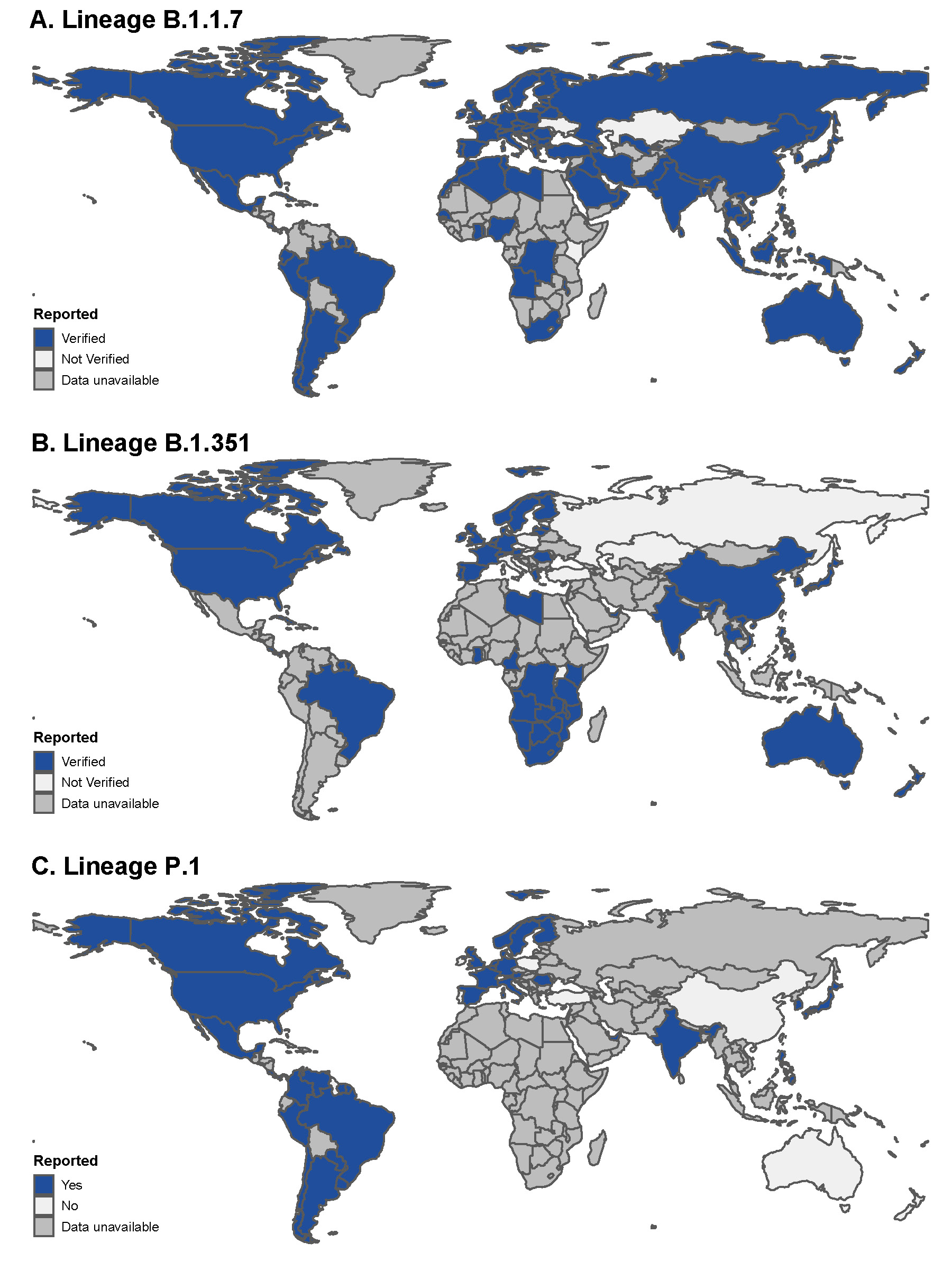

#### Fig S2. The fold change of neutralizing antibodies against variants compared to reference strains among non-variant-infected individuals based on matched samples.

Neutralizing antibodies were determined in live virus neutralization assay (purple dot), lentivirus-vector pseudovirus neutralization assay (green dot), and VSV-vector pseudovirus neutralization assay (blue dot). Each connection line represents a paired sample.

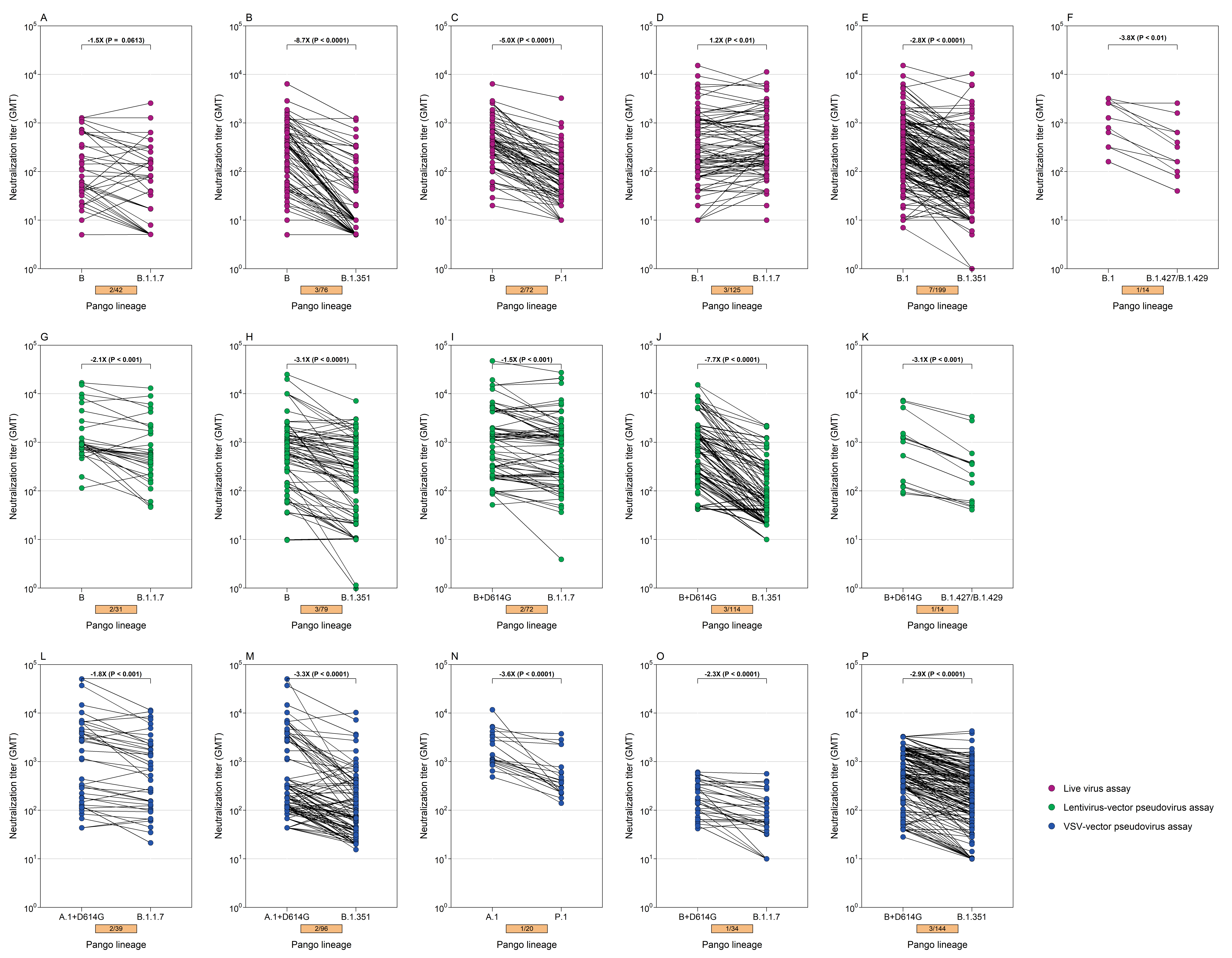

#### Fig S3. Neutralizing antibodies against SARS-CoV-2 variants in variant-infected individuals by neutralization assays

Neutralizing antibodies of individuals infected with reference strains (blue dot), lineage B.1.1.7 (red dot), and B.1.351 (green dot) against same lineages were determined in **A)** live virus neutralization assay and **B)** lentivirus-vector pseudovirus neutralization assay. The solid point represents the GMT and the error bar represents the 95% confidence interval. The scattering dot represents individual titers.

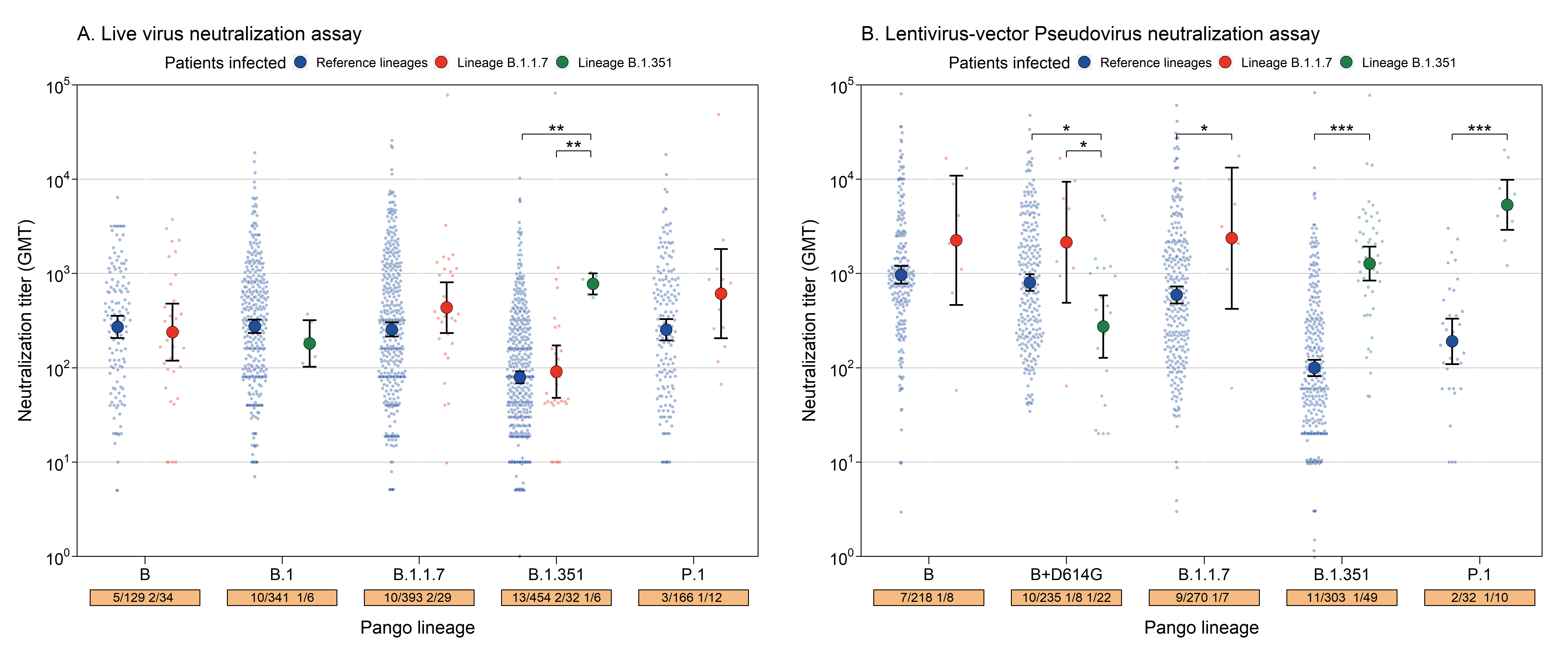

#### Fig S4. The fold change of neutralizing antibodies against variants compared to reference strains among uninfected vaccine recipients in live virus neutralization assay.

The subtitles represent vaccine subjects after the dose of different vaccine platforms. Each connection line represents a paired sample.

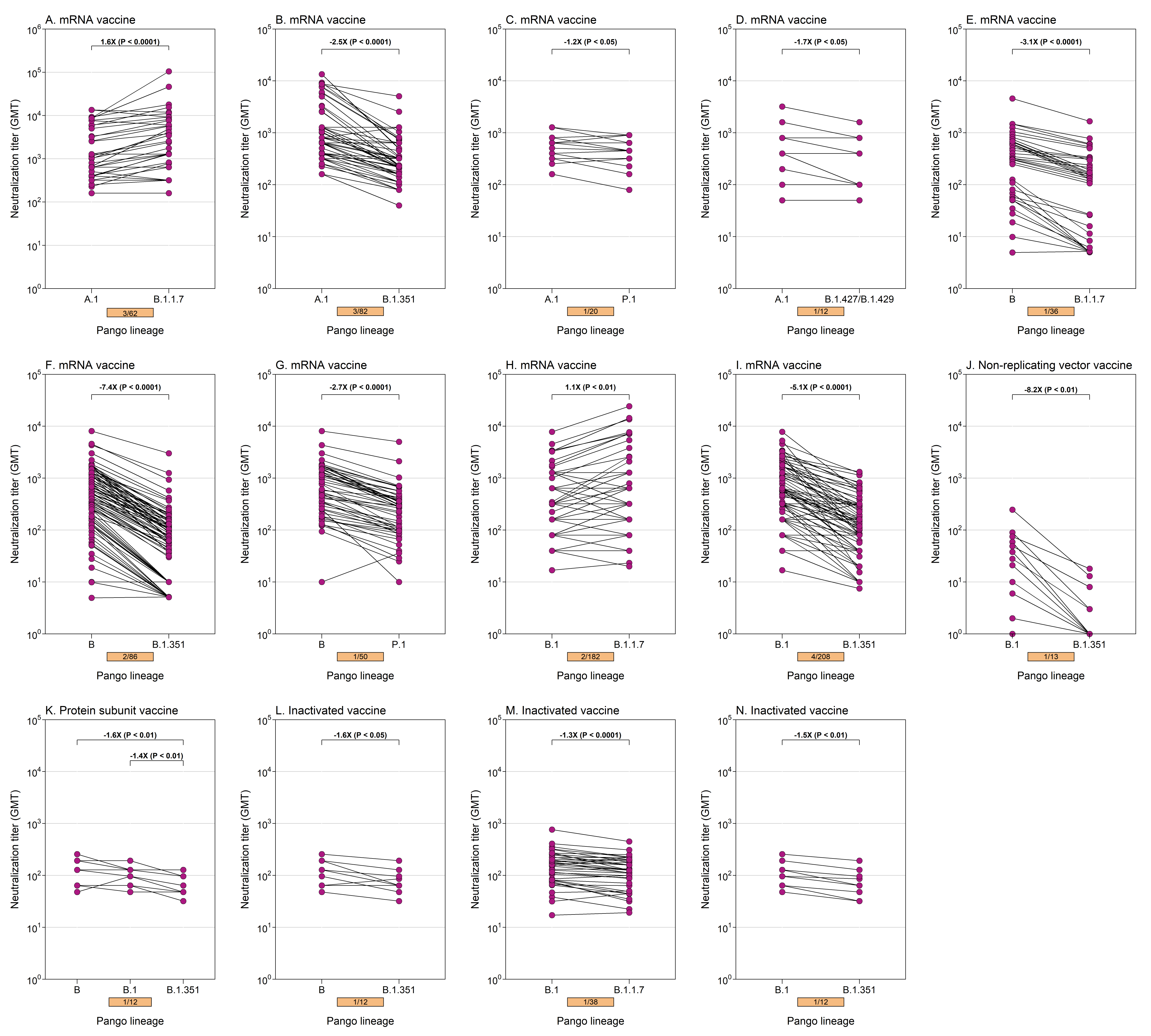

#### Fig S5. The fold change of neutralizing antibodies against variants compared to reference strains among uninfected vaccine recipients in lentivirus-vector pseudovirus neutralization assay.

The subtitles represent vaccine subjects after the dose of different vaccine platforms. Each connection line represents a paired sample. Each connection line represents a paired sample.

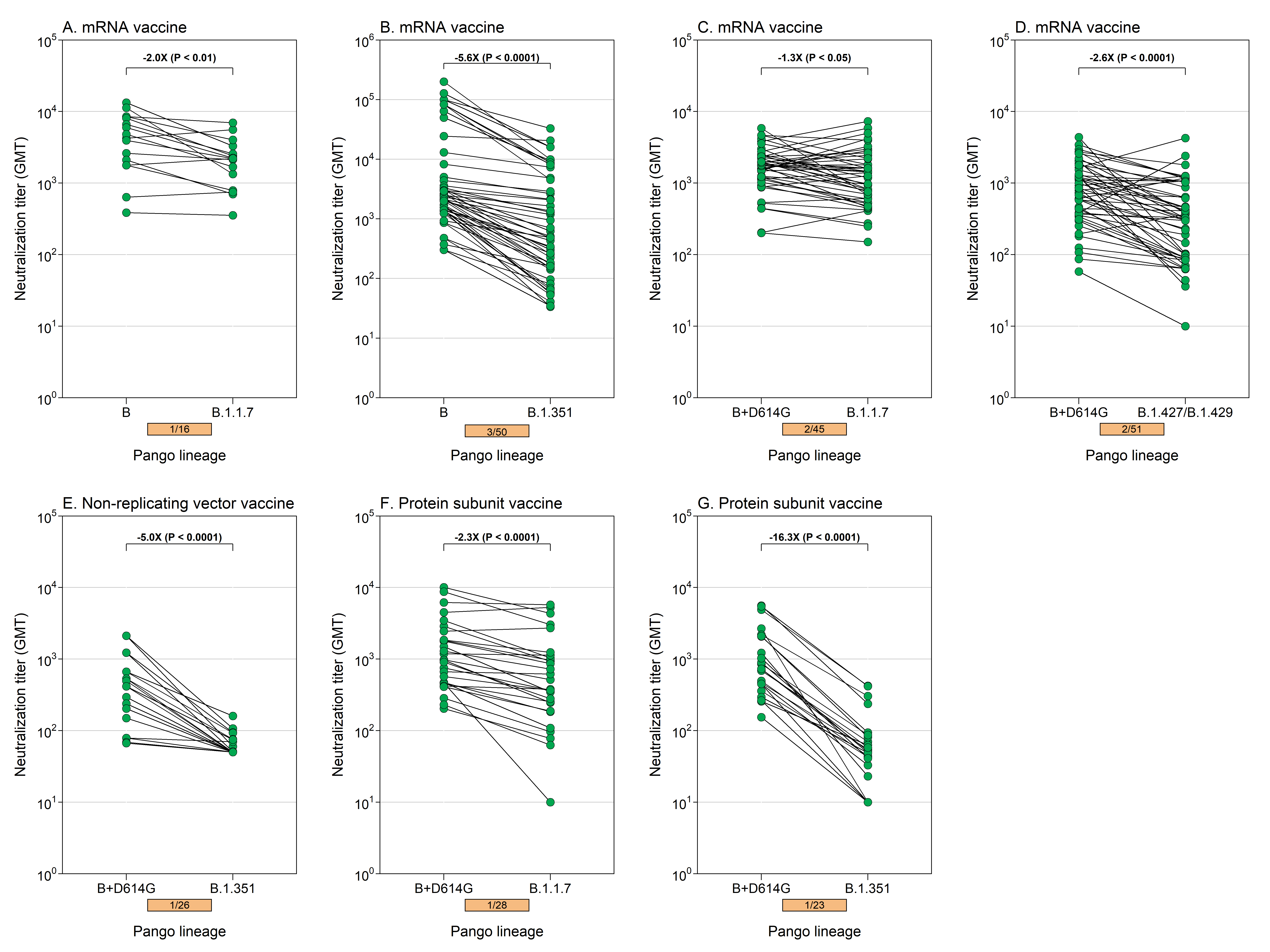

#### Fig S6. The fold change of neutralizing antibodies against variants compared to reference strains among uninfected vaccine recipients in VSV-vector pseudovirus neutralization assay.

The subtitles represent vaccine subjects after the dose of different vaccine platforms. Each connection line represents a paired sample. Each connection line represents a paired sample.

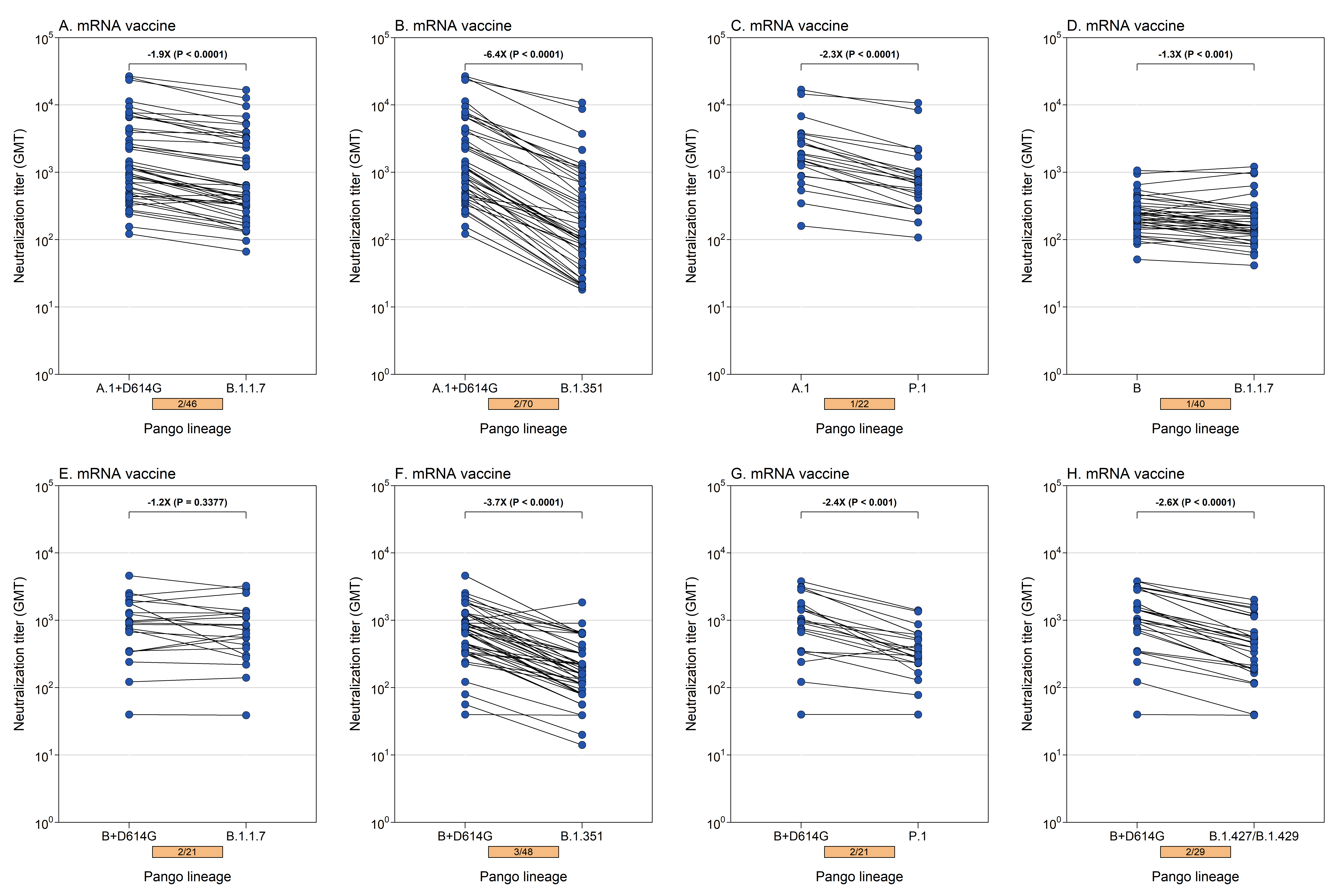

#### Fig S7. Comparison of Neutralizing antibodies against SARS-CoV-2 variants between previously-infected and uninfected vaccine recipients by neutralization assays and vaccine platforms

**A)** live virus neutralization assay, **B)** lentivirus-vector pseudovirus neutralization assay, **C)** VSV-vector pseudovirus neutralization assay. The solid point represents the GMT and the error bar represents the 95% confidence interval. The scattering dot represents individual titers. N-R vector, non-replicating vector.

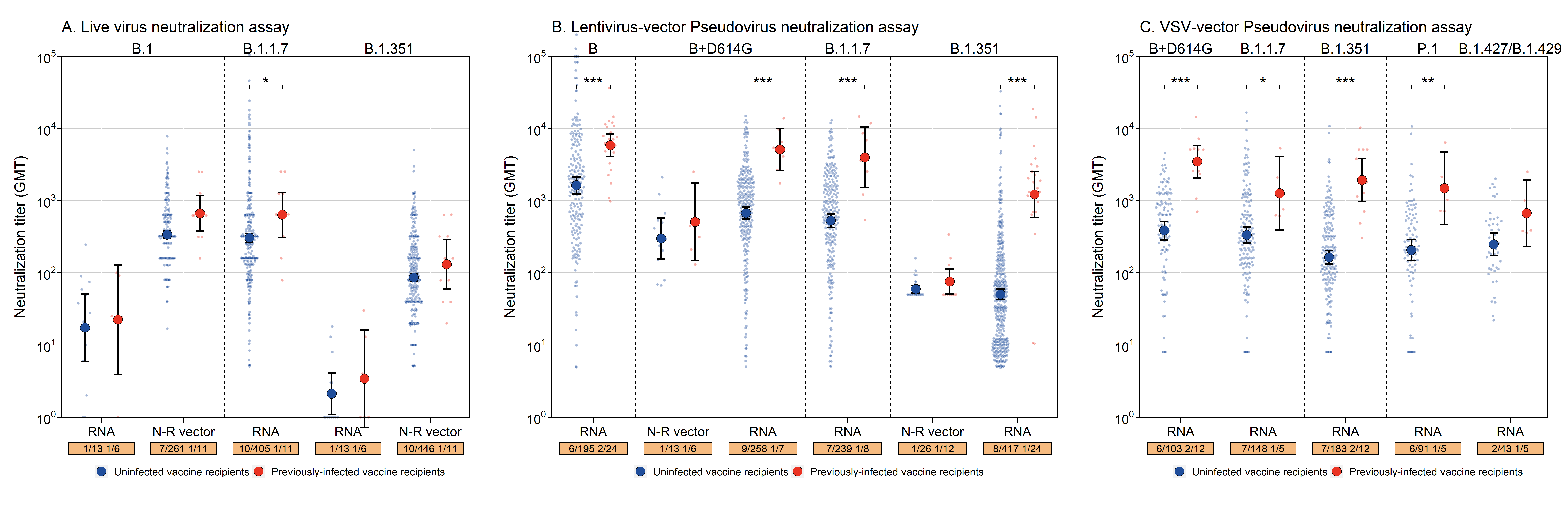

#### Fig S8. Comparison between GMT of neutralizing antibodies induced by natural infections and vaccination

**A)** live virus neutralization assay, **B)** lentivirus-vector pseudovirus neutralization assay, **C)** VSV-vector pseudovirus neutralization assay. The solid point represents the GMT and the error bar represents the 95% confidence interval. The scattering dot represents individual titers.

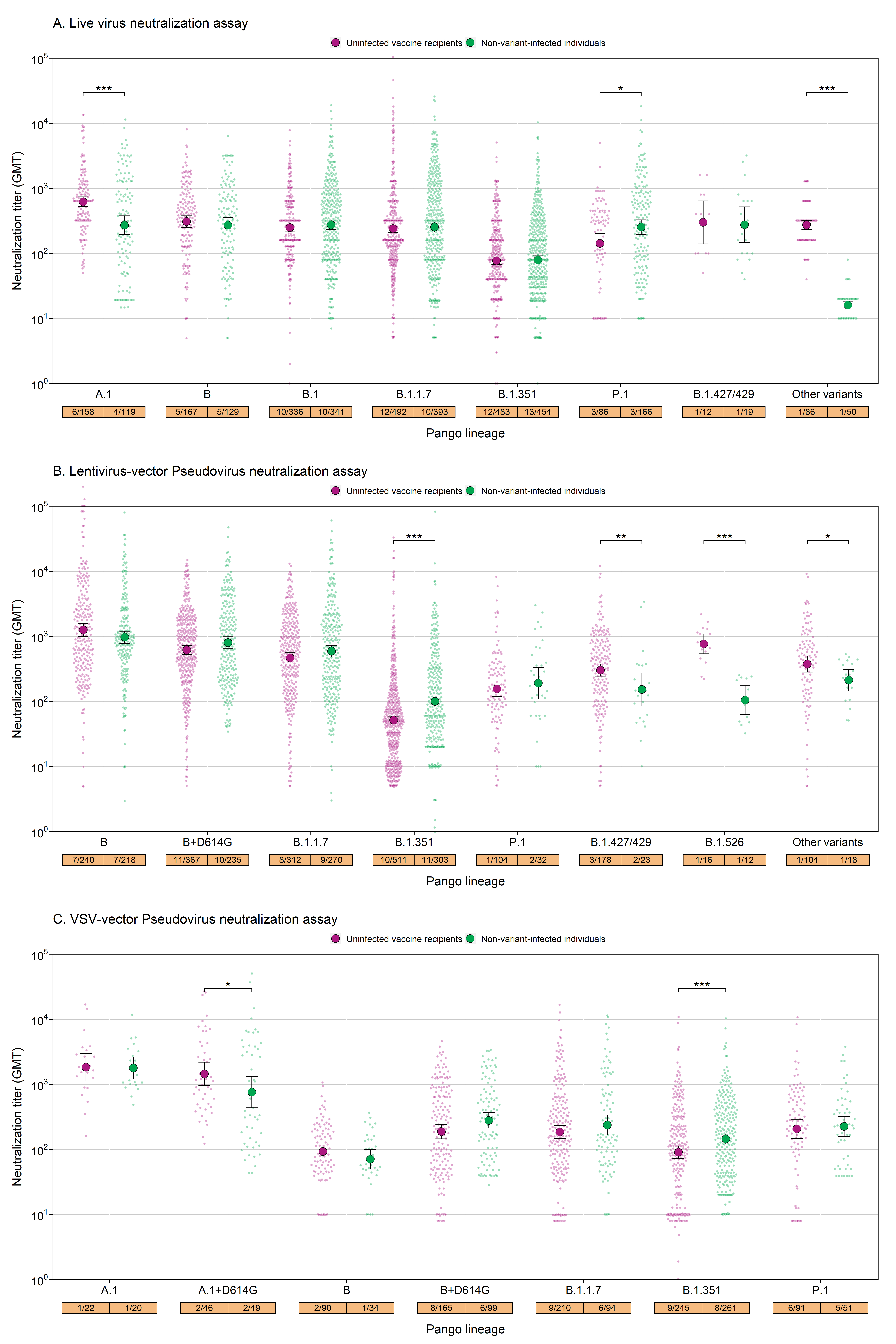

#### Fig S9. The dynamic of neutralizing antibodies by sampling interval among non-variant-infected patients in live virus neutralization assays

Distribution of sampling interval was done against **A)** lineage B and B.1, **B)** B.1.1.7, and **C)** B.1.351. The sampling time grouping is not very strict here.

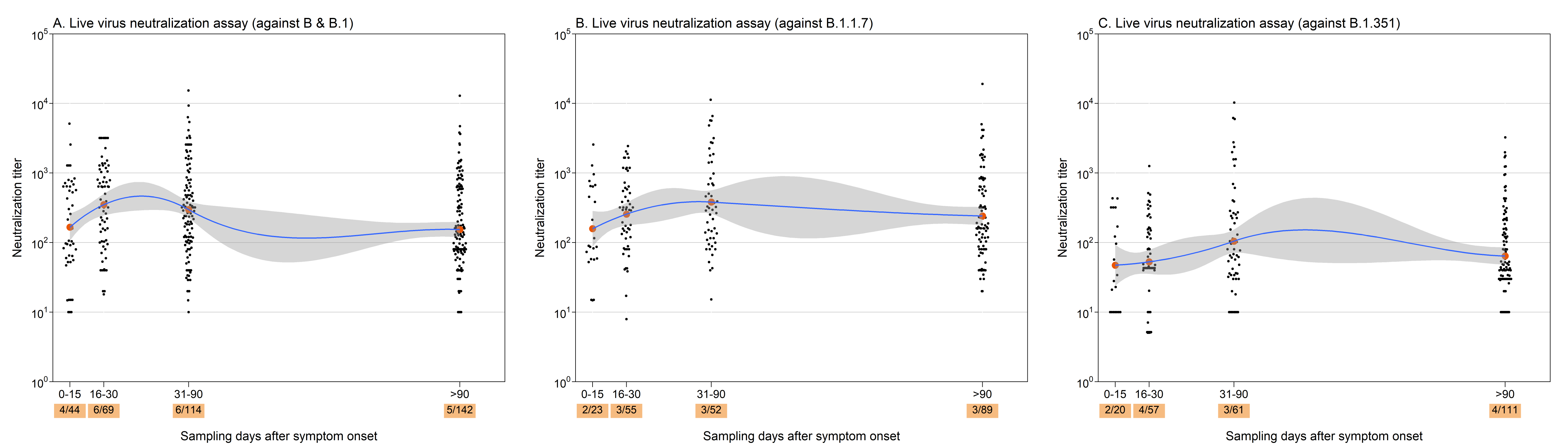

#### Fig S10. Subgroup analysis of neutralizing antibodies among non-variant-infected individuals by clinical severity based on two neutralization assays

Subgroup analysis of clinical severity was based on **A)** live virus neutralization assay and **B)** lentivirus-vector pseudovirus neutralization assay.

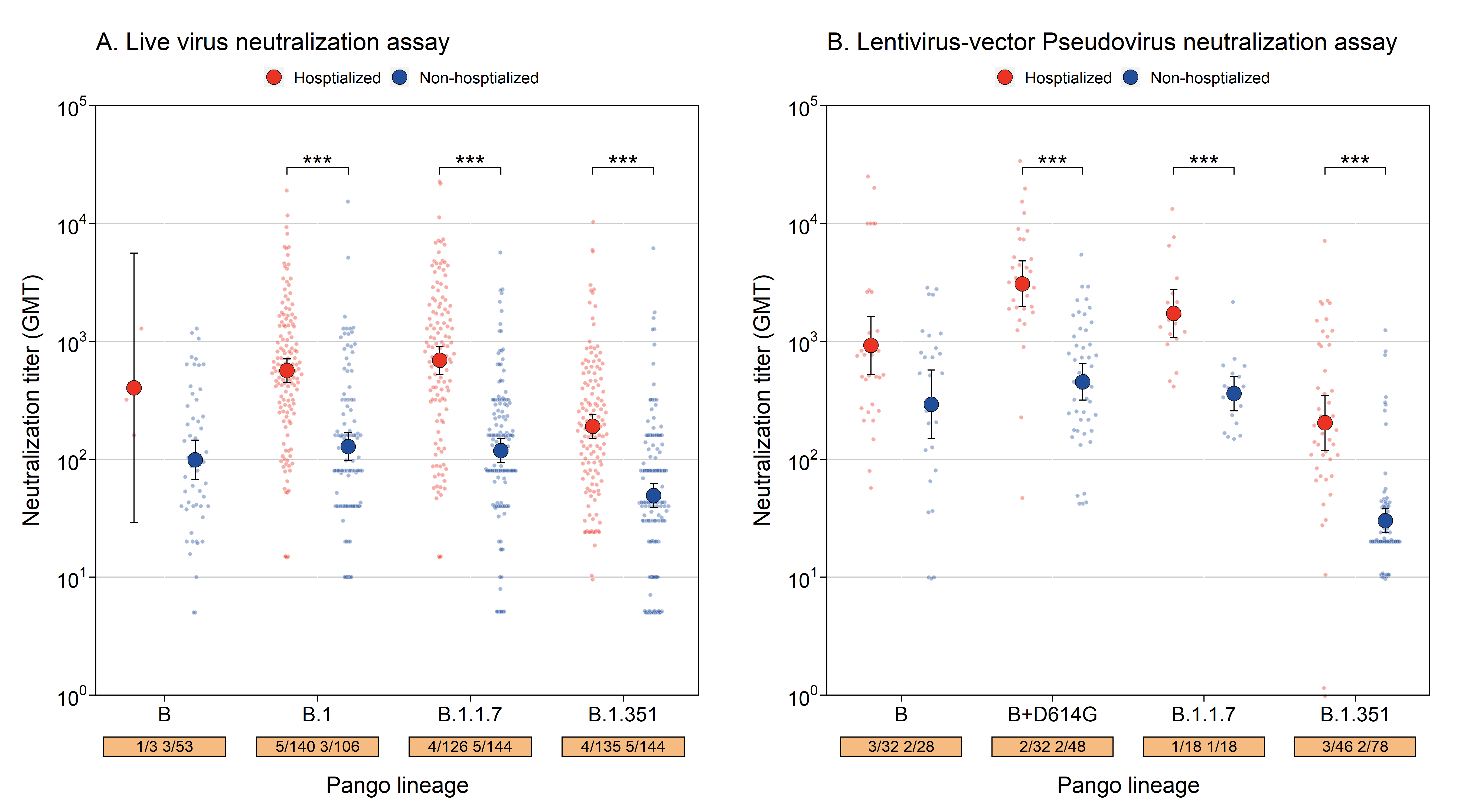

19.	Hu J, Peng P, Wang K, et al. Emerging SARS-CoV-2 variants reduce neutralization sensitivity to convalescent sera and monoclonal antibodies. *Cellular & molecular immunology*.

20.	Li R, Ma XC, Deng JY, et al. Differential efficiencies to neutralize the novel mutants B.1.1.7 and 501Y.V2 by collected sera from convalescent COVID-19 patients and RBD nanoparticle-vaccinated rhesus macaques. *Cellular & molecular immunology*.

35.	Wibmer CK, Ayres F, Hermanus T, et al. SARS-CoV-2 501Y.V2 escapes neutralization by South African COVID-19 donor plasma. *Nature Medicine*.

42.	Zhou H, Dcosta B, Samanovic M, Mulligan M, Landau N, Tada T. B.1.526 SARS-CoV-2 variants identified in New York City are neutralized by vaccine-elicited and therapeutic monoclonal antibodies. bioRxiv; 2021.

59.	Creech CB, Walker SC, Samuels RJ. SARS-CoV-2 Vaccines. *JAMA* 2021.

60.	Doria-Rose N, Suthar MS, Makowski M, et al. Antibody Persistence through 6 Months after the Second Dose of mRNA-1273 Vaccine for Covid-19. 2021.

61.	World Health Organization. COVID-19 Weekly Epidemiological Update. 2021. <https://www.who.int/emergencies/diseases/novel-coronavirus-2019/situation-reports> (accessed Mar 26 2021).
